# A coordinate-to-coordinate machine learning framework for contrast-agnostic brain target localization

**DOI:** 10.64898/2026.11.24.26368764

**Authors:** Alaa Taha, Jaime Thrower, Dhananjhay Bansal, Arun Thurairajah, Mohamad Abbass, Mackenzie Snyder, Joy Zhao, Ezgi Coskun, Violet M. Liu, Greydon Gilmore, Ali R. Khan, Jonathan C. Lau

## Abstract

Representing a structure by a coordinate can be challenging particularly for structures which lack conspicuity, whether due to their small size, lack of contrast, or image artifact. In this paper, we developed a machine learning framework to localize brain targets from the coordinates of salient brain landmarks (i.e., coordinate-to-coordinate) that is agnostic to image contrast. We make use of a validated brain landmark annotation protocol inspired by classical stereotactic methods and a dataset which spans a wide range of MRI field strengths and neurodegenerative diseases. We apply this framework to predict deep brain stimulation targets, achieving an accuracy of 1-1.5 mm validated against expert rater annotations on ultra-high field (UHF) MRI. Results demonstrated comparable localization accuracy on test-retest data acquired using both standard and UHF MRI. Our proposed framework outperformed deformable registration on clinical MRI and performed comparably on multispectral UHF MRI. We deployed our model in the form of a website (stereotaxy.afids.io) and openly release all coordinate data. This framework is agnostic to MRI contrast and offers a method for rapid coordinate localization with applications more broadly in neuroimaging as well as for specialized settings like surgical planning.

## 1. Introduction

The origins of coordinate-based brain mapping can be traced back to Horsley and Clarke (Horsley & Clarke, 1908), who designed a stereotactic apparatus using Cartesian coordinates to localize brain structures relative to cranial landmarks. Building on this foundation, Jean Talairach (Schaltenbrand et al., 1977; Talairach et al., 1957) leveraged anatomical landmarks, such as the anterior commissure (AC) and posterior commissure (PC), for more refined brain structure localization (i.e., the proportional grid normalization). These early innovations laid the groundwork for population-based brain templates that emerged with the advent of magnetic resonance imaging (MRI). For example, the MNI305 template (Collins et al., 1994) aligned 305 individual MRI scans using AC and PC, overcoming the idiosyncrasies of using a single subject brain as a template. Subsequent advances in deformable registration further refined these templates, culminating in modern standards such as the MNI152 space (Fonov et al., 2009), now widely employed for aligning anatomical data. Despite these advances, localizing small, poorly visualized structures with millimetric accuracy remain challenging, particularly when direct anatomical visualization is limited by image resolution, contrast, or artifacts.

Deep brain stimulation (DBS) is a stereotactic procedure involving the surgical implantation of electrodes in specific brain structures. It is well established for the management of a wide array of movement disorders such as Parkinson’s disease (PD), essential tremor (ET), dystonia, and Tourette’s syndrome, as well as for select psychiatric conditions including obsessive-compulsive disorder and treatment-resistant depression (Lozano et al., 2019). Small deviations in electrode position during DBS of the subthalamic nucleus (STN), as an example, result in side-effects including paresthesias, pyramidal symptoms, diplopia, and dysarthria (Buhmann et al., 2017). Meanwhile, suboptimal placement of the electrode by a few millimeters (mm) can lead to reduced clinical outcomes (Anheim et al., 2008) and also put patients at risk for post-operative cognitive decline (Reich et al., 2022). Thus, preoperative localization of DBS targets must approach millimeter-level accuracy for optimal therapeutic benefit.

Achieving the level of accuracy needed in DBS is complicated by the limitations of clinical neuroimaging. Most MRI scanners used in clinical settings operate at 1.5 or 3 Tesla (T), which may not always provide the needed resolution for clear visualization of all DBS targets. Furthermore, patient motion (exacerbated in individuals with movement disorders), suboptimal MRI protocols, and geometric distortions can further degrade image quality (Boutet et al., 2021; Chandran et al., 2016; Lau et al., 2018).

Ultra-high field (UHF; ≥ 7 Tesla) MRI enables clear delineation of small brain regions, since the main magnetic field strength results in enhanced signal-to-noise ratio and tissue contrast that can be exploited to improve spatial resolution (Abosch et al., 2010; Duchin et al., 2012; Lau et al., 2017, 2020; Lenglet et al., 2012). However, UHF-MRI systems are not commonly available, posing an obstacle to the widespread use in clinic (Clarke et al., 2020; Middlebrooks et al., 2025). As a result, determining the optimal DBS target still benefits from indirect localization techniques that rely on predefined coordinate spaces.

Existing methods for indirect localization can be grouped into two categories: (1) atlas-based consensus coordinates and (2) atlas-based registration. Atlas-based coordinate approaches describe consensus coordinates of a target relative to the midpoint along the AC-PC line, known as the mid-commissural point (MCP). The MCP is susceptible to intersubject variability, as no single atlas-based consensus coordinate perfectly captures the anatomical differences observed on various populations. Meanwhile, atlas-based (i.e., deformable) registration techniques estimate the deformation field used to propagate atlas labels from a stereotactic space (e.g., MNI) to individual brains. Deformable registration yields errors on the order of 1–5 mm (Abbass et al., 2022; Lau et al., 2019; Miller et al., 2023), with lower quality clinical scans prone to failing thus requiring manual intervention. Neurosurgical planning typically uses gadolinium-enhanced T1-weighted MRI (MRI-gad) as the reference image. However, most atlases are derived from data without gadolinium enhancement, so there is no direct one-to-one mapping. This adds complexity to registration that may behave unpredictably (Abbass et al., 2022, 2025) and limits the ability to integrate standard neuroimaging workflows (Ogunsanya et al., 2024; Warntjes et al., 2014).

Machine learning (ML) models for automatic localization of brain structures have the potential to offer a faster and more generalizable alternative to deformable registration (Baniasadi et al., 2023; Ren et al., 2025). However, only a limited number of studies have evaluated and adapted ML frameworks to DBS targeting while achieving generalizability on clinical (e.g., MRI-gad) scans (Baniasadi et al., 2023). Furthermore, due to the intensive process of acquiring manual ground-truth labels, many approaches have focused on matching accuracy of well-curated parameters during deformable registration approaches (i.e., training labels generated from quality controlled deformable registration segmentation data) (Baniasadi et al., 2023).

In this study, we validate a coordinate-to-coordinate ML framework to accurately localize brain structures solely from x, y, z coordinates of surrounding anatomical landmarks (see Figure 1). Our target localization approach is dependent only on validated anatomical landmarks (Abbass et al., 2022; Lau et al., 2019), which are reproducible on various MRI field strengths and modalities (Taha et al., 2023). This framework was designed with generalizability and explainability in mind, such that it can be applied to other brain targets while enabling understanding of relationships between predefined landmarks and the targeted brain region, including structures targeted in DBS (e.g., the subthalamic nucleus [STN] and caudal zona incerta [cZI]). Our framework integrates well within the clinical workflow, which already involves preoperative localization of landmarks (e.g., AC and PC). Central to our approach is an open framework, which involves the release of our coordinate data, code, and model in the form of a website (“stereotaxy.afids.io”).

**Figure 1.**
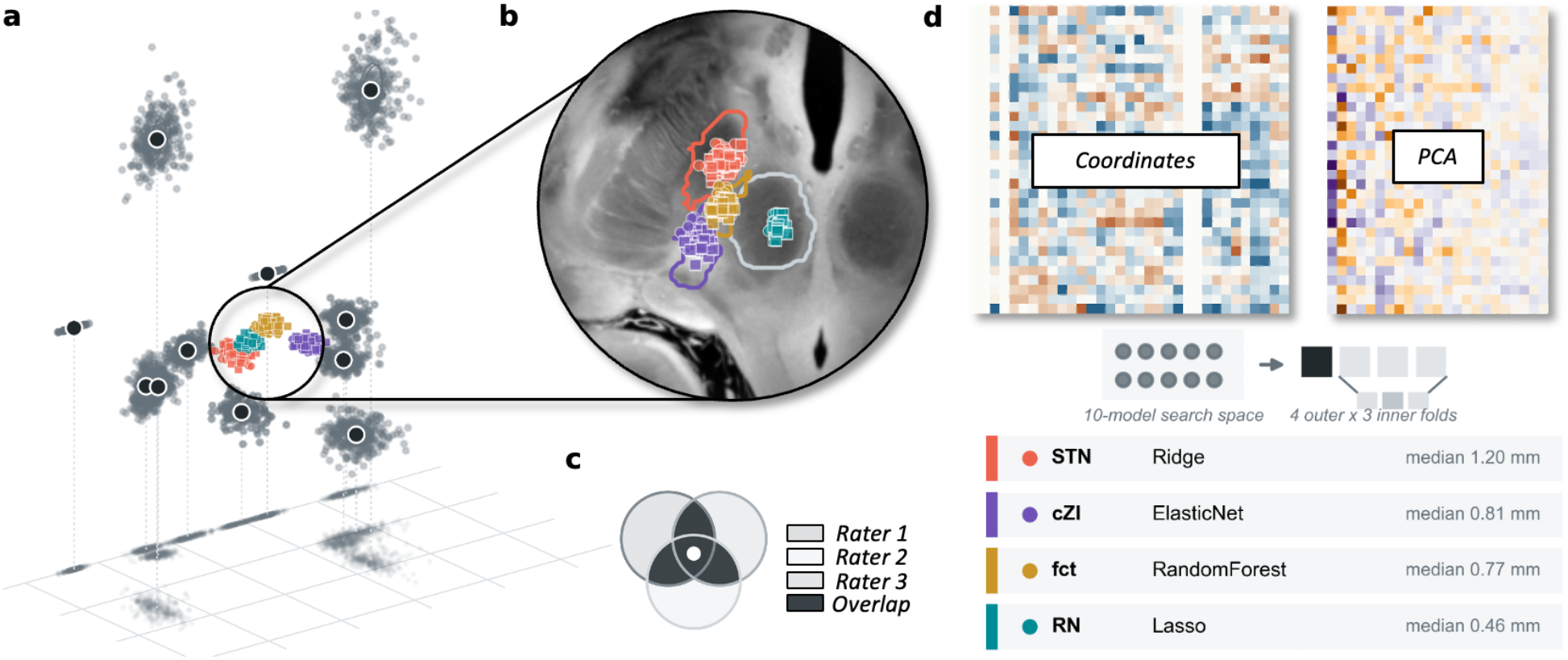
Coordinate-to-coordinate framework for brain target localization. **(a)** Anatomical fiducial (AFID) coordinates after AC-PC alignment, centering on the mid-commissural point (MCP), and left-to-right reflection, shown as a three-dimensional point cloud. Black circles with white outlines represent the mean AFID location; colored points are the ground-truth centroids of the four targets: STN (coral), cZI (purple), fct (gold), and RN (teal). **(b)** The circled target cluster is magnified on an axial slice of the 7T ex vivo human brain, with outlines of the target segmentations. **(c)** Ground-truth definition: each target is segmented by at least two raters (gray) on 7T MRI, combined by majority vote, and represented by its center of mass. **(d)** Learning pipeline: AFID coordinates are standardized and reduced by PCA, ten regression architectures are screened under nested cross-validation, with results showing the architecture with the best performance

## 2. Methods

### 2.1 Participants

We leverage 202 MRI scans on which we have 6,464 ground truth anatomical landmarks applied in accordance with a validated annotation protocol (Lau et al., 2019) by more than 20 human raters. Below is a breakdown of this data (see Table 1):

**Table 1.**
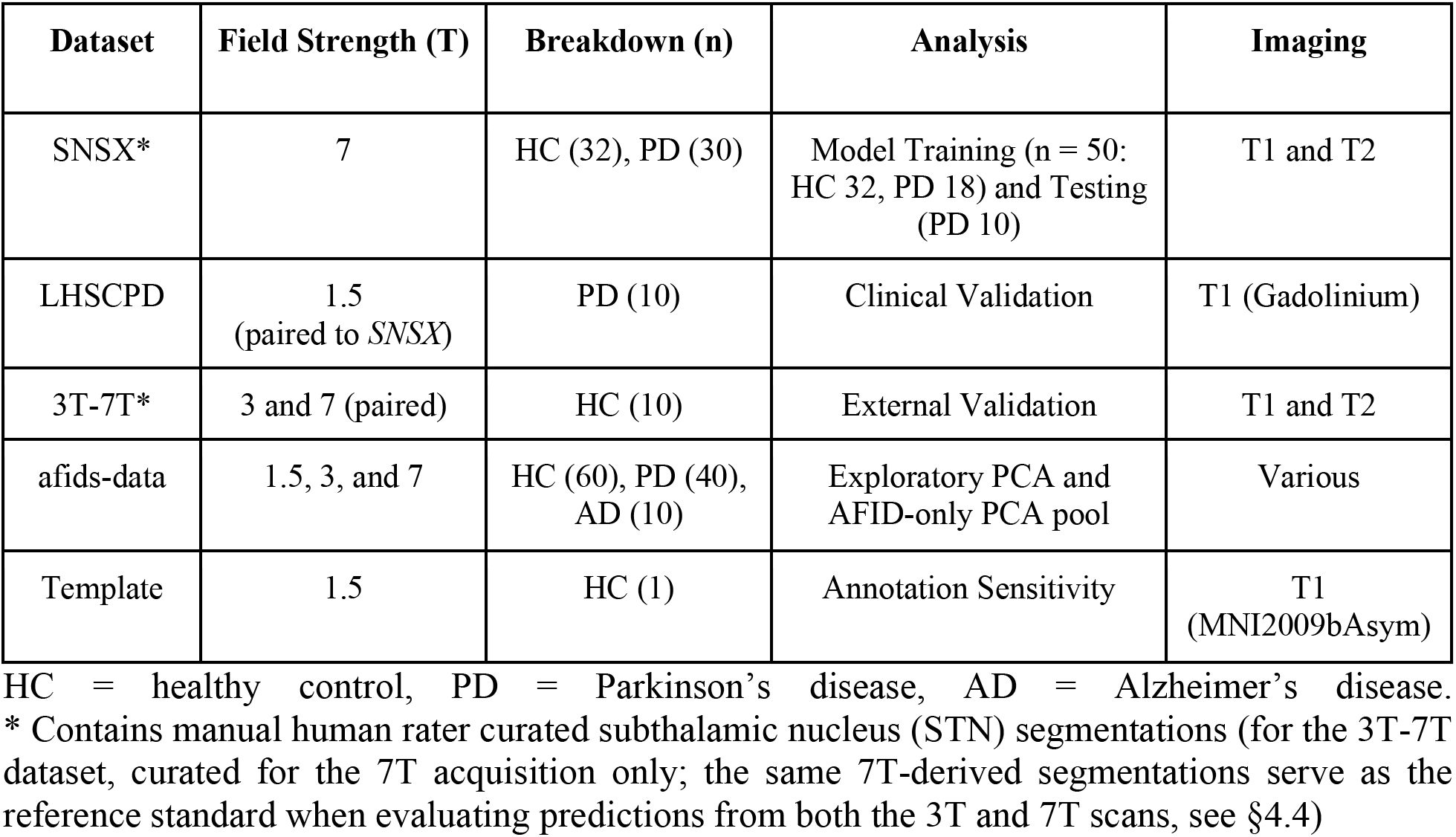
Various datasets leveraged as part of this study, all of which have curated anatomical landmark annotations.

*The Stereotactic Neurosurgery (SNSX) dataset.* This dataset consists of 62 participants (32 healthy controls and 30 scheduled for DBS). Participants were imaged using a 7T head-only MRI scanner (Siemens Magnetom; Siemens Healthineers, Erlangen, Germany) at the Centre for Functional and Metabolic Mapping (CFMM) in Western University (REB# 109045). The MRI sequences used for this study were: 1) 3D T1w MP2RAGE and 2) 3D optimized T2w fast-spin echo (T2 SPACE).

*The London Health Sciences Centre Parkinson’s Disease (LHSCPD) dataset.* This dataset consists of a subset (n = 10) of the same participants as in the SNSX dataset, scheduled to undergo DBS at London Health Sciences Centre (LHSC; REB# R-17-156). Participants were imaged using a 1.5T MRI scanner (Signa, 1.5T, General Electric, Milwaukee, Wisconsin, USA). The MRI sequence used in this study was a 3D T1w MP2RAGE with gadolinium contrast (termed “MRI-gad” in subsequent sections).

*An openly released 3T and 7T (3T-7T) dataset (Chen et al., 2023)*, consisting of healthy participants (n = 10) imaged using the following sequences: 1) 3D T1w MP2RAGE and 2) 3D T2 SPACE. Participants are imaged at both 3T and 7T (i.e., test-retest).

*A subset of a previously released (afids-data) dataset (Taha et al., 2023)*, which aggregated brain landmark annotations (n = 110) on various populations (healthy, Parkinson’s disease, and Alzheimer’s disease [AD]). This dataset is independent of the previously mentioned datasets.

A single T1-weighted MNI152NLin2009bAsym and 7T ex vivo template scan (Fonov et al., 2009; Edlow et al., 2019), carrying AFIDs ground-truth annotations, was additionally used to evaluate the sensitivity of STN predictions to AFID annotation variability.

### 2.2 Coordinate Data Annotation

We employ a previously validated protocol for the annotation of anatomical fiducials (AFIDs; https://afids.github.io/afids-protocol) providing a point-based sampling of multiple brain structures (Lau et al., 2019). In this study, we selected a subset of 16 AFIDs (see Supplementary Figure S1) described by the protocol which (1) circumscribe the midbrain and surrounding structures (e.g., DBS targets) and (2) can be localized within 1.5 mm by expert and novice human raters (Abbass et al., 2022).

#### AFID Annotations

All annotations were performed in accordance with the AFIDs protocol via 3D Slicer 4.10 (Fedorov et al., 2012) Rater placements per AFID were averaged to curate the ground-truth coordinates (x, y, and z).

#### DBS Target Annotations

All DBS target annotations were performed via 3D Slicer (v.4.10) using the smallest configuration of the paint brush (1 mm) and the pencil drawing tool on 7T T2w imaging. Rater segmentations per subject were selected via majority voting (>50%) and then collapsed to a center of mass constituting the ground truth coordinates (see Lau et al., 2019 for more details). For this work, we annotate the STN, cZI, fasciculus cerebellothalamicus (fct), and red nucleus (RN).

Each scan was annotated by at least two human raters (see Supplementary Table S1 for a breakdown of rater demographic data). The ground truth coordinates were quality controlled (QC) by three raters (AT, JZ, and EC). We provide QC files (*.html format; see Supplementary Attachment A) with accompanying code and ground truth coordinates (*.fcsv) used in this work. Supplementary Attachment B provides an interactive 3D-visualization of all the coordinates and datasets.

### 2.3 Coordinate Data Processing

An AC-PC alignment was performed followed by centering coordinates at the MCP. Coordinates on the left hemisphere were reflected to the right by inverting the sign of x-coordinate after AC-PC transformation and MCP centering. Thus, each participant constituted two augmented examples. Preliminary analysis using training data was conducted to investigate linear correlations between coordinates and to characterize the variance structure of the concatenated x, y, z AFID coordinates via PCA (Figure 2b–d); this exploratory PCA is descriptive and was not itself used for model training. For ML model training and evaluation, the PCA was instead refit independently within each training partition (see Section 2.4), retaining the top PCs capturing 99% of variance in that partition each time.

**Figure 2.**
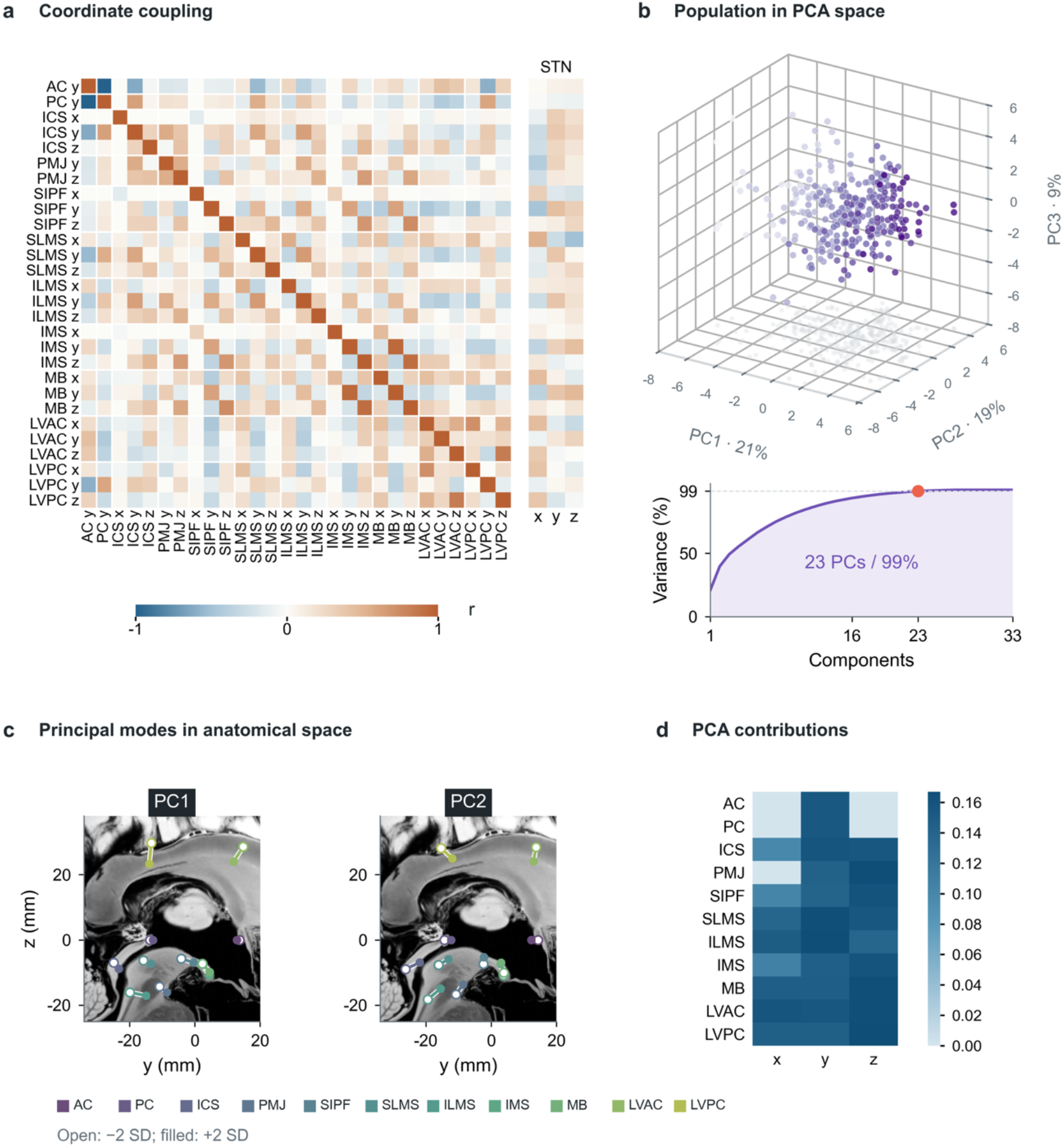
Correlation of landmarks and principal component analysis (PCA). **(a)** Pearson correlations for 28 AFID coordinate features, grouped by landmark, and their correlations with right-STN x, y, and z (right columns). AFID–AFID correlations use 320 hemisphere examples; AFID–STN correlations use 47 STN-labelled training participants retained after outlier processing. **(b)** Development examples projected onto the first three standardized principal components (color encodes PC1 score); below, cumulative explained variance reaches 99% at 23 components. **(c)** Landmark configurations reconstructed at -2 SD (open) and +2 SD (filled) along PC1 and PC2, with other component scores set to zero, shown in the y-z plane on a sagittal slice of the 7T ex vivo brain; segments connect the same landmark. **(d)** Contribution of each landmark coordinate to the PCA, summed over components as absolute loading multiplied by explained-variance ratio. These values describe variance structure, not supervised predictive importance (see Supplementary Figure S6).

### 2.4 Data Split and Machine Learning

AFID and STN coordinate data of participants from the SNSX dataset were used for ML training (50 participants: 32 HC and 18 PD). Ten further participants from the SNSX dataset (all PD) were assigned to a reserved testing dataset used solely to report the accuracy of our model. These participants were also the same ones from the LHSCPD dataset enabling evaluation of our model on clinical quality 1.5T MRI-gad scans. Finally, the *3T-7T* dataset was used for external validation to report prediction accuracy at 3T and 7T MRI.

To select the regression architecture, we screened ten candidate models covering linear, kernel-based, and ensemble methods: Ridge regression (Hoerl & Kennard, 1970), BayesianRidge, ElasticNet, Lasso, partial least squares (PLS), support vector regression with a radial basis function kernel (SVR), k-nearest neighbors (KNN), random forest, a multi-layer perceptron (MLP), and Extreme Gradient Boosting (XGBoost; Chen & Guestrin, 2016), an ensemble method building on the gradient boosting framework (Friedman, 2001) that can model non-linear relationships. All candidate architectures used the same preprocessing pipeline, with a separate model fitted for each of the x, y, and z coordinates, and hyperparameters were tuned within each training fold using model-specific searches (Supplementary Methods S1.3 and S2). For Ridge, the regularization strength was tuned over a logarithmic grid (α = 10⁻³ to 10⁶, 200 log-spaced values; the deployed model was tuned over a wider grid, 10⁻¹⁰⁰ to 10¹⁰⁰, and its selected values, α = 4.6–11.6, lie inside both ranges). Candidates were compared on cross-validated ED (Supplementary Table S2; Supplementary Figure S2). To examine the trade-off between model simplicity and complexity, we additionally compared Ridge (the simplest regularized linear candidate) with XGBoost (the most flexible non-linear candidate) on per-axis errors from the same cross-validation (Supplementary Figure S3).

ML models were trained via a nested cross-validation (CV) strategy to tune hyperparameters and evaluate performance (scikit-learn; Pedregosa et al., 2011). The outer CV loop involved a 4-fold split of all training data (repeated five times with different participant-to-fold assignments) for evaluating and comparing the candidate models and the inner CV loop involved a 3-fold split CV strategy for tuning hyperparameters. To prevent data leakage, we were intentional in our handling of input data, specifically: (1) the folds were curated such that for a given training example (i.e., participant) the left and right hemisphere target labels (e.g., STN) were grouped together (Supplementary Methods S2), preventing left and right labels from appearing separately on training and testing; (2) furthermore, group-based preprocessing (i.e., PCA) was performed independently within each fold. PCA was fit on each fold’s training partition augmented with an additional pool of AFID-only scans (the 110 afids-data scans and the two SNSX participants without STN annotations, 112 scans; disjoint from all train/test subjects) for consistency with the deployed model; this retained 24 components at the 99%-variance threshold. Outer-loop errors were used only for model comparison; the selected model was then refit on all training data and evaluated once on the reserved testing datasets (section 3.3; Table 2). We compute the x, y, z mean squared error (MSE) and Euclidean distance (ED) to evaluate models.

**Table 2.** Model Mean Squared Error (MSE) and Euclidean distance (ED) performance on paired datasets. Main-text summary statistics (§3.3) pool all four rows below (80 STNs total: Testing Data [SNSX], Clinical Validation [LHSCPD], External Validation [3T], and External Validation [7T]); each row here also reports its own 20-STN cohort separately. ±Values are median [IQR] (top) and mean ± SD (bottom). MSE values are in mm²; ED values are in mm.

| **Dataset** | **MSE** |  |  | **ED** | **Wilcoxon Signed Rank Test**<br>(α = 0.05/4) |
|---|---|---|---|---|---|
|  | **x** | **y** | **z** |  |  |
| Testing Data (SNSX) | 0.48 [0.22–0.88]<br>0.59 ± 0.49 | 0.30 [0.08–0.76]<br>0.59 ± 0.77 | 0.43 [0.03–0.73]<br>0.43 ± 0.38 | 1.19 [0.98–1.50]<br>1.22 ± 0.37 | *MSE x: p* = 0.13 (ns)<br>*MSE y: p* = 0.09 (ns)<br>*MSE z: p* = 0.11 (ns)<br>*ED: p* = 0.12 (ns) |
| Clinical Validation (LHSCPD) | 0.24 [0.09–0.68]<br>0.49 ± 0.58 | 0.80 [0.08–0.93]<br>0.74 ± 0.87 | 0.57 [0.11–0.79]<br>0.60 ± 0.58 | 1.32 [1.08–1.58]<br>1.29 ± 0.42 |  |
| External Validation (3T) | 0.10 [0.05–0.44]<br>0.39 ± 0.57 | 0.43 [0.06–1.30]<br>0.68 ± 0.76 | 0.07 [0.02–0.65]<br>0.48 ± 0.78 | 1.20 [0.80–1.49]<br>1.15 ± 0.49 | *MSE x: p = 0.22 (ns)*<br>*MSE y: p = 0.31 (ns)*<br>*MSE z: p = 0.52 (ns)*<br>*ED: p = 0.15 (ns)* |
| External Validation (7T) | 0.07 [0.02–0.27]<br>0.34 ± 0.60 | 0.26 [0.04–0.93]<br>0.61 ± 0.78 | 0.16 [0.03–0.49]<br>0.42 ± 0.57 | 1.08 [0.70–1.40]<br>1.07 ± 0.48 |  |

### 2.5 Model Robustness and Comparative Evaluation

#### Generalization to Other Imaging Modalities

To investigate model accuracy on clinical imaging, STN coordinate predictions on 1.5T MRI-gad scans were statistically compared to paired ground-truth counterpart 7T MRI-nogad scans. To further demonstrate robustness on both research imaging and external data, we also evaluated model predictions on the 3T-7T dataset. Statistical comparisons were in AC-PC space, and two independent Wilcoxon signed-rank tests were performed across x,y,z MSE and ED with Bonferroni correction (α = 0.05/4). Normality was assessed with the Shapiro-Wilk test; because squared per-axis errors were strongly right-skewed (p < 0.001), all comparisons used non-parametric Wilcoxon signed-rank tests and are summarized as median [IQR] alongside mean ± SD.

#### Benchmarking

We performed statistical comparisons between our model and deformable registration for STN localization using our 1.5T MRI-gad dataset (i.e., LHSCPD) and paired 7T-MRI counterpart (i.e., SNSX). The Distal atlas segmentations in MNI space (Chakravarty et al., 2006; Ewert et al., 2018) were warped to native space using presets from a nonlinear deformation framework validated via 11,000 nonlinear warps on more than 100 subjects (Ewert et al., 2019). We applied this framework as implemented in Lead-DBS software (v.3.1; Neudorfer et al., 2023). Groups were compared using a Wilcoxon signed-rank test.

#### Robustness to Annotation Variability

To investigate model dependency on AFID localization error, we evaluated STN prediction accuracy using 38 rater annotations in a common reference space (i.e., MNI). For this analysis, the AFID protocol was applied independently by 10 human raters.

#### Landmark Prediction Generalizability

To evaluate whether the coordinate-to-coordinate approach generalizes beyond STN localization, we performed a leave-one-AFID-out analysis using the 16-AFID subset from section 2.2. For each AFID, we treated its coordinates as the prediction target and used the remaining landmarks as predictors. We applied the same participant-grouped nested cross-validation pipeline from Section 2.4. We restricted the comparison (see Supplementary Figure S4) to the nine landmarks that define the coordinate space for midbrain and subthalamic targets.

### 2.6 Extending This Framework to Other Neuromodulation Targets

Having established the accuracy and robustness of the coordinate-to-coordinate framework for STN localization, we next tested whether the same approach would generalize, extending the framework to three additional midbrain structures relevant to DBS planning either as known surgical targets or adjacent features of interest: the caudal zona incerta (cZI), fasciculus cerebellothalamicus (fct), and red nucleus (RN). A single annotation pass therefore would have the potential to support prediction of all four targets addressed in this study (STN, cZI, fct, and RN). The ten candidate regression models described earlier were evaluated at each target, using the same shared preprocessing pipeline, candidate-model roster, and statistical-comparison methodology as the STN model screen (Section 2.4; see Supplementary Methods S1), within a Leave-One-Group-Out Cross-Validation (LOGO-CV) framework on the SNSX healthy-control training cohort (32 participants and 32 folds). Given the proof-of-concept scope of this extension and the absence of paired multi-modality or deformable-registration ground truth for these targets, evaluation is restricted to LOGO-CV within the training cohort. Full methodological details are provided in Supplementary Methods S1.

### 2.7 Model Deployment

Building on the previous open software infrastructure of the AFID project, we deployed our model on a website (“stereotaxy.afids.io”). This enables users to upload their AFID placements as specified by our modified protocol (see Supplementary Figure S1) and receive target coordinate predictions alongside an AC-PC transformation matrix. AFIDs are uploaded in native image space; the website derives the AC-PC/MCP transformation from them, predicts in that space, and returns the STN prediction together with the transformation matrix.

## 3. Results

### 3.1 Rater Curated Data

Demographic data for human raters involved in annotations are made available in Supplementary Table S1. The overall localization error on the subset of 16 AFIDs used in our framework was 0.84 ± 0.31 mm, and rater STN segmentation overlap was 0.78 ± 0.03 (Dice similarity coefficient). Right and left STN segmentations did not differ and were combined in subsequent summaries. Median STN volumes were 136.68 mm³ [IQR 126.43–147.37] in the healthy control (HC) cohort and 126.28 mm³ [IQR 104.69–147.92] in the Parkinson’s disease (PD) cohort (p = 0.048; Mann-Whitney U test). Median STN centroid coordinates in MCP space for the HC cohort were x = 10.05 [IQR 9.72, 10.70], y = −0.90 [IQR −1.39, −0.40] and z = −3.27 [IQR −3.52, −2.83] mm, and for the PD cohort were x = 10.29 [IQR 9.32, 10.87], y = −1.02 [IQR −1.31, −0.48] and z = −3.65 [IQR −4.14, −3.05] mm (x: p = 0.93, y: p = 0.91, z: p = 0.002; Mann-Whitney U test). After Bonferroni correction (α = 0.05/4), only the z-coordinate difference remained significant.

### 3.2 Preliminary Analysis and Principal Component Analysis (PCA)

Pairwise correlations were computed for the x, y, and z coordinates of 16 AFIDs, reduced to 11 unique landmark positions after the hemisphere-mirroring step described in section 2.3 (33 features total; Figure 2a). PCA was employed to reduce the dimensionality of features used for ML model training. In this exploratory, training-only PCA, the top 23 principal components (PCs) captured 99% of the variation in the data (see Figure 2b); the production pipeline, fit with the additional AFID-only pool, retained 24 (Section 2.4). Further exploration of AFID contributions to PCs revealed relatively broad AFID contributions (see Figure 2d). Strong linear associations between AFID coordinates and STN x, y, and z coordinates were observed (Pearson’s r = −0.50 to 0.52) in the training data (Figure 2a), supporting the use of a linear regression framework.

### 3.3 Machine Learning Model Performance

#### Model Training

Across the 10-model screen (one model per x, y, z coordinate; nested CV), ridge regression ranked first by median ED (0.85 mm; BayesianRidge had a marginally lower mean ED, 0.95 vs. 0.96 mm). After Holm-Bonferroni correction, Ridge was not statistically distinguishable from BayesianRidge, PLS, RandomForest, or ElasticNet, but significantly outperformed SVR, Lasso, XGBoost, KNN, and MLP (Supplementary Table S2; Supplementary Figure S2). In the follow-up comparison with XGBoost, Ridge had lower error (participant-level median ED 0.89 vs. 0.96 mm; Wilcoxon signed-rank test, Bonferroni-corrected p = 0.002; lower error for 72% of participants), with significantly lower x and y but not z errors (Supplementary Figure S3). Given its top rank and its simplicity, Ridge was selected as the final model for predicting STN coordinates in this study.

#### Model Testing

Across all testing data (80 STNs), the mean ± SD squared error was 0.45 ± 0.56, 0.66 ± 0.79, and 0.48 ± 0.59 mm² along x, y, and z. The overall median ED was 1.20 mm (IQR: 0.84–1.53 mm; participant-level bootstrap 95% CI for the median: 1.01–1.38 mm; mean ± SD: 1.18 ± 0.44 mm), and 98.8% of STN predictions (79 of 80) were within 2 mm (Table 2). Held-out error was insensitive to the PCA variance threshold (median ED 1.22–1.29 mm for thresholds from 80% to 99.9%) and decreased with increasing training-set size (from 1.48 mm with 8 to 1.22 mm with 50 training participants; Supplementary Figure S5). Cross-validated error was higher in PD than in HC training participants (median ED 1.13 vs. 0.80 mm; Mann-Whitney U test, p = 0.003; Supplementary Results S3), consistent with the higher error on the all-PD held-out test set than in cross-validation.

### 3.4 Model Generalizability and Comparisons

#### Validation for Other Imaging Modalities

We evaluated model performance on different imaging modalities by analyzing test-retest data acquired at various field strengths (see Figure 3a–c). Ten participants with both 1.5T MRI-gad (*LHSCPD*) and 7T MRI-nogad (*SNSX*), as well as ten participants with both 3T and 7T MRI (*3T-7T*), were included (see Section 2.4). There were no significant differences in x, y, z MSE and ED on the two paired imaging modalities.

**Figure 3.**
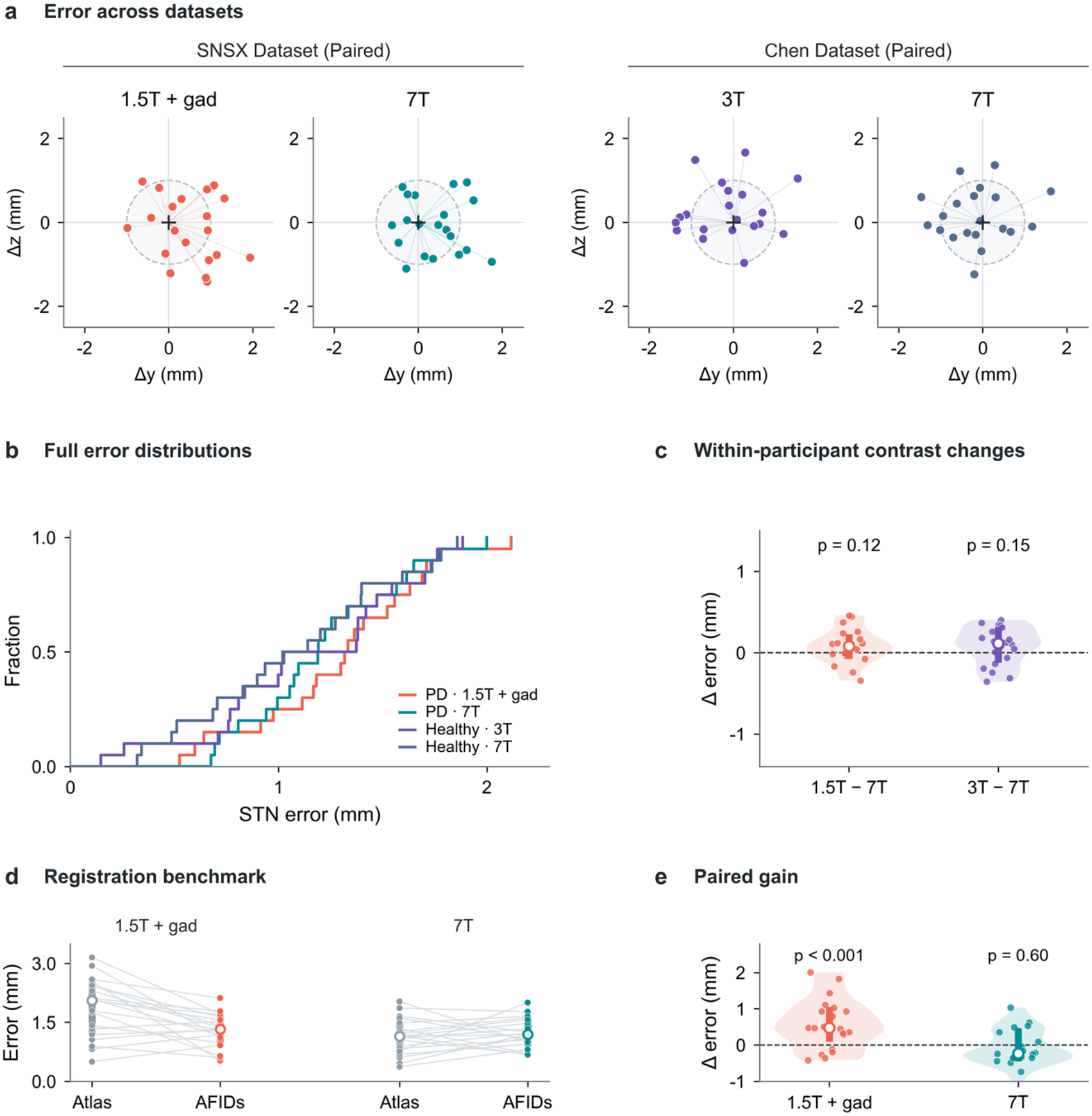
Held-out STN localization across acquisitions and against registration. **(a)** Prediction residuals relative to each hemisphere’s reference STN location, projected onto the y–z plane, for the clinical pair (1.5T with gadolinium and 7T; LHSCPD/SNSX dataset) and the external pair (3T and 7T; 3T-7T or Chen dataset). The dashed circle marks a projected distance of 1 mm. **(b)** Empirical cumulative distributions of three-dimensional Euclidean error for the four acquisitions. **(c)** Within-participant differences in Euclidean error (1.5T-7T and 3T-7T). Points are paired hemispheres, shaded shapes kernel density estimates, thick bars interquartile ranges, and open circles medians; p values are paired Wilcoxon signed-rank tests (1.5T vs. 7T, p = 0.12; 3T vs. 7T, p = 0.15). **(d)** Paired Euclidean errors of atlas-based deformable registration (ANTs/Lead-DBS) and AFID-based prediction on the clinical 1.5T gadolinium-enhanced and 7T acquisitions (n = 20 hemispheres each); gray lines connect matched hemispheres. **(e)** Registration error minus AFID error (positive values favour the AFID model); paired Wilcoxon signed-rank tests, 1.5T with gadolinium p < 0.001, 7T p = 0.60.

#### Model Comparison and Performance Benchmarking

We benchmarked our model against deformable registration (see Figure 3d–e). On 1.5T MRI-gad scans, our model statistically outperformed registration (Wilcoxon signed-rank test, n = 20 hemispheres from 10 patients, p < 0.001). Conversely, on multispectral 7T MRI-nogad data (i.e., registration warps computed using both T1 and T2 imaging), no significant difference in performance was observed between registration and our model (p = 0.60).

#### Robustness to Annotation Variability

To assess the influence of landmark annotation error on model performance, we examined the relationship between rater localization error and prediction error. A moderate correlation was observed (*Pearson r* = 0.47; p < 0.001; see Figure 4a–b).

**Figure 4.**
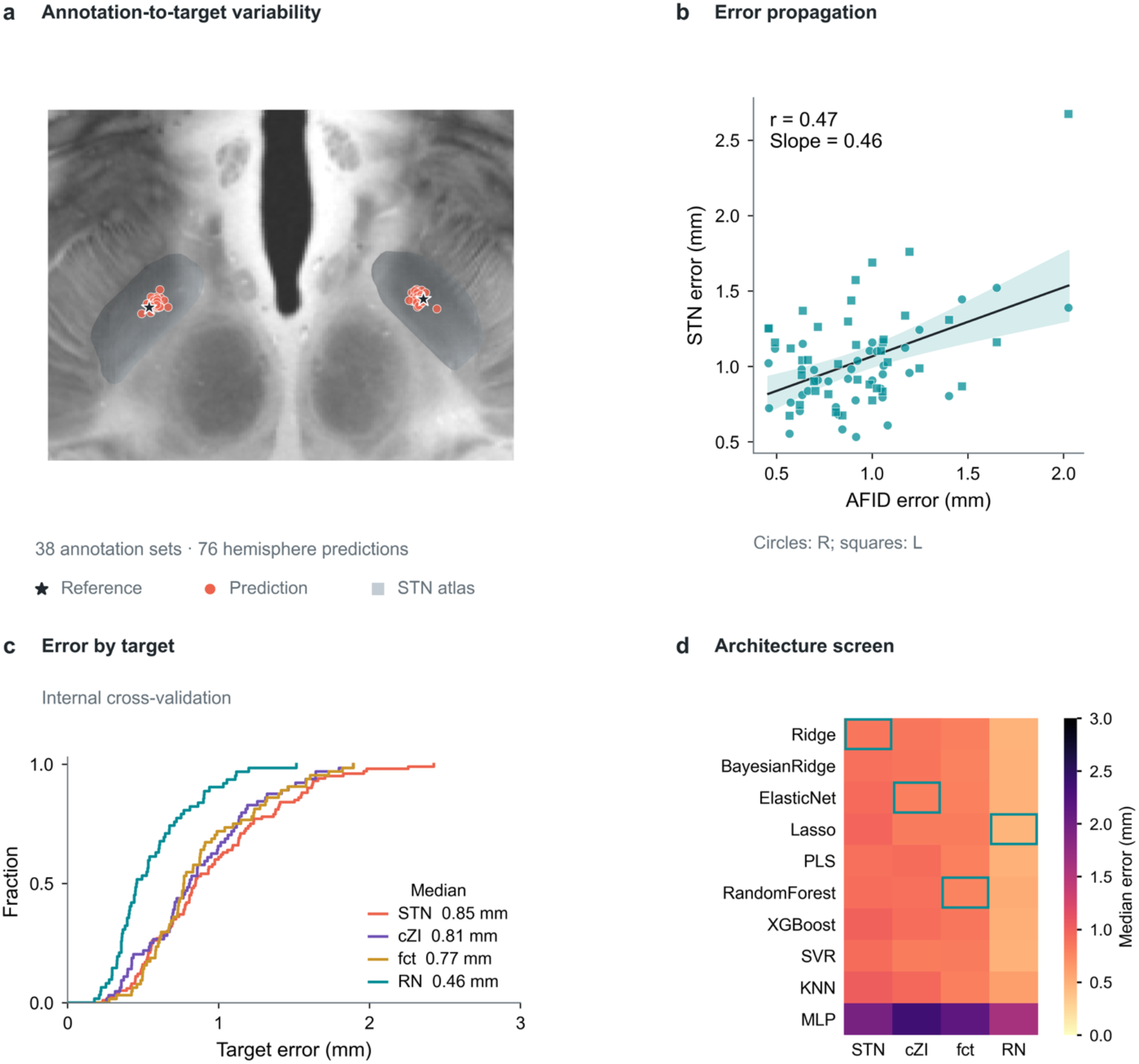
Robustness to annotation variability and extension to additional targets. **(a)** STN predictions from 38 independent AFID annotations by 10 raters on the MNI152NLin2009bAsym template (76 hemisphere predictions; coral points) and the reference STN coordinates (stars), overlaid on an axial slice of the 7T ex vivo brain with the STN atlas (gray; DISTAL). **(b)** Mean AFID annotation error versus STN prediction error (circles, right; squares, left hemisphere; Pearson r = 0.47, p < 0.001; ordinary least-squares slope = 0.46). The shaded band is the 95% confidence interval of the fitted mean and does not account for repeated raters and hemispheres. **(c)** Empirical cumulative error distributions under cross-validation for the selected architecture of each target. **(d)** Median cross-validated error of all candidate architectures for each target; outlined cells mark the lowest median per target. STN is shown for reference (5 × 4-fold cross-validation); cZI, fct, and RN use leave-one-participant-out cross-validation for both model selection and error estimation.

#### Generalization to other Midbrain Landmarks

To test whether the coordinate-to-coordinate framework would generalize beyond STN prediction, we performed a leave-one-AFID-out analysis for nine other midbrain landmarks (see section 2.5; Supplementary Figure S4); errors are per-hemisphere Euclidean distances averaged over five repeats of participant-grouped 4-fold cross-validation (50 participants). AC and PC were localized almost exactly (median ED 0.06 mm; IQR 0.04–0.10) and their errors are identical, since in AC-PC/MCP space each is determined by the other (together they define the AC-PC axis and share a single degree of freedom); they therefore do not test the framework and were excluded from the pooled summary below. Of the remaining seven landmarks, MB (median ED 0.67 mm; IQR 0.47–0.87), IMS (0.68 mm; IQR 0.48–0.93), and SIPF (0.79 mm; IQR 0.54–1.04) were reconstructed most accurately, followed by ICS (1.17 mm; IQR 0.72–1.69), SLMS (1.28 mm; IQR 0.80–1.74), and ILMS (1.38 mm; IQR 1.10–1.84), with PMJ (1.64 mm; IQR 1.08–2.30) the least accurate. Pooled over these seven landmarks, the overall median ED was 1.00 mm (IQR: 0.67–1.53 mm; 700 hemisphere estimates), comparable in magnitude to the inter-rater localization variability previously reported for direct manual AFID annotation (0.36–1.68 mm; Lau et al., 2019). These results indicate that the ridge-regression coordinate-to-coordinate approach is not specific to STN localization and generalizes to reconstructing other midbrain landmarks from the surrounding AFID configuration.

### 3.5 Generalizability to other deep brain structures

To assess the broader applicability of our framework, we applied the coordinate-to-coordinate approach, including the same model-architecture screen used for the primary target (section 3.3), using an identical candidate roster and preprocessing pipeline (see Supplementary Methods S1). Model benchmarking identified the best-performing architecture for each target: ElasticNet for cZI, RandomForest for fct, and Lasso for RN, though for cZI and fct most other candidates were not statistically distinguishable from the winner after Holm-Bonferroni correction (only KNN and MLP for cZI, and only MLP for fct, were significantly worse), whereas for RN Lasso significantly outperformed Ridge, BayesianRidge, ElasticNet, and MLP but was not distinguishable from PLS, SVR, XGBoost, RandomForest, or KNN (Supplementary Tables S3–S5; Supplementary Figure S2). On LOGO-CV, these top performing models achieved a median ED of 0.81 mm (IQR: 0.56–1.12; mean ± SD: 0.87 ± 0.40), 0.77 mm (IQR: 0.61–1.11; 0.89 ± 0.38), and 0.46 mm (IQR: 0.36–0.70; 0.56 ± 0.28) for the cZI, fct, and RN, respectively (Figure 4c). Complete comparison tables for all 10 candidate models are provided in Supplementary Tables S3 (cZI), S4 (fct), and S5 (RN), with median errors for all architectures in Figure 4d and the corresponding cross-validation error distributions in Supplementary Figure S2.

## 4. Discussion

We demonstrated that coordinates of predefined brain landmarks can be leveraged to predict the coordinates of other surrounding brain regions with millimetric accuracy. By employing a coordinate-to-coordinate ML framework, our model is agnostic to imaging modality and resolution. Furthermore, we demonstrated that our model outperforms conventional methods when using less optimal clinical scans (e.g., 1.5T MRI-gad) while performing comparably when using multispectral research scans (i.e., 7T MRI with both T1 and T2 contrasts). Finally, we packaged this model in the form of a website, enabling its use in research or clinical contexts.

### 4.1 Rater Curated Data

#### AFID Annotations

Previous AFID validation studies demonstrated millimetric localization in healthy (Lau et al., 2019) and clinical populations (Abbass et al., 2022). The subset of 16 AFIDs in our ML framework was selected for their spatial proximity to the STN, sampling the more predictable morphological features of the midbrain region. Although the complete AFIDs protocol (i.e., 32 AFIDs) typically requires 20-40 minutes following rater training (Lau et al., 2019), we anticipate a substantial reduction in annotation time due to the reduced number of AFIDs used here. To facilitate reproducibility and completeness, we provide a protocol tailored to the 16 AFIDs used in this work, illustrated at true anatomical position on the MNI152NLin2009bAsym template (see Supplementary Attachment C and Supplementary Figure S1).

#### STN Annotations

Inter-rater agreement for STN segmentation was strong, with a Dice similarity coefficient of 0.78 ± 0.03. This is consistent with prior manual segmentation studies of the STN and nearby subcortical structures (Camlidag et al., 2014; Miller et al., 2023). Prior work also highlighted that no statistically significant differences were found between left and right STN annotations (Duchin et al., 2018; Miller et al., 2023), justifying their combination in subsequent analyses. On average, the STN volume was reduced in the PD cohort relative to controls (128.03 ± 29.67 mm³ and 136.56 ± 16.24 mm³, *p* = 0.047). However, this volume difference did not remain statistically significant after Bonferroni correction. Prior studies reported conflicting findings, with some showing no significant difference in STN volume between PD and HC (Alkemade et al., 2017; Camlidag et al., 2014), while others reported a reduction in the PD cohort (Colpan & Slavin, 2010; Patriat et al., 2020). We believe a meta-analytic approach is needed for more conclusive findings. Analysis of STN coordinates relative to the MCP revealed a significant inferior shift (i.e., z-axis) in PD patients compared to controls (*p* = 0.002). This finding may support prior work reporting region-specific morphological changes associated with PD (Kaya et al., 2019). Interestingly, previous work comparing STN coordinates between PD and essential tremor patients reported no significant differences (Duchin et al., 2018); however, essential tremor may not serve as an appropriate baseline for comparison.

### 4.2 Preliminary Analysis and Principal Component Analysis (PCA)

Our analysis demonstrates that brainstem AFIDs exhibit strong internal spatial correlations, consistent with prior reports of geometrical regularity in this region (Perera et al., 2024). Structures such as the superior lateral mesencephalic sulcus (SLMS), superior interpeduncular fossa (SIPF), and mammillary bodies (MB) were tightly coupled along shared axes, suggesting conserved spatial organization driven by developmental and structural constraints (Párraga et al., 2016). PCA supported this anatomical consistency, with the top five components capturing over 60% of the variance in AFID coordinates. This concentration of variance in a modest number of components reflects meaningful redundancy in the AFID configuration and supports the dimensionality reduction used for downstream ML applications, though the remaining components (23 total for 99% of variance; section 3.2) still contribute non-trivial information preserved for model training. Importantly, variance was broadly distributed over AFIDs rather than dominated by a few landmarks, reflecting coordinated spatial variation across regions. When correlating AFIDs with STN coordinates, SLMS ranked among the top predictors for all axes (strongest |r| per STN axis = 0.38–0.52). The MB and SIPF also demonstrated strong associations (MB: |r| = 0.38–0.47; SIPF: |r| = 0.32–0.48). Univariate correlation does not, however, equal contribution to a multivariate model. In a supervised analysis (Supplementary Figure S6), removing any single landmark changed cross-validated error by at most 0.07 mm, with the largest increases for LVPC, PMJ, LVAC, and SLMS, and nested landmark selection with 4-11 landmarks gave similar error (median ED 0.85–0.90 mm); the ventricular landmarks and PMJ thus contributed as much as SLMS despite weaker univariate correlations, consistent with redundancy among AFIDs. Taken together, these results support a coordinate-based localization approach, leveraging conserved anatomical relationships within the midbrain and diencephalic regions to infer the position of brain targets like the STN.

### 4.3 Machine Learning Model Performance

Our model achieved a median localization error of 1.20 mm (IQR: 0.84–1.53 mm; bootstrap 95% CI for the median: 1.01–1.38 mm; mean ± SD: 1.18 ± 0.44 mm) on all testing data. This performance is comparable to the localization variability of AFIDs themselves, previously reported to range from 0.36 to 1.68 mm (Lau et al., 2019). For context, a recent study reported a mean inter-rater centroid localization error of 1.9 ± 1.07 mm between two expert annotators (Miller et al., 2023). A similar coordinate-to-coordinate framework (Engelhardt et al., 2021) achieved a mean error of 1.33 ± 1.64 mm; however, direct comparison is limited, as their target was the active contact of a DBS electrode rather than an anatomically defined structure. Notably, Guo et al. (2007) benchmarked six STN targeting methods and found that a combined anatomical and functional approach yielded a localization error of 1.70 ± 0.7 mm. Clinically, errors under 2 mm are conventionally considered within the range of acceptable (Kremer et al., 2023), particularly when considering the role of intraoperative neurophysiology conventionally to optimize targeting as well as the advantages of modern DBS electrode designs that allow for current steering. A recent multicenter study similarly explored AI-assisted STN-DBS surgical planning using a supervised statistical atlas-based approach, reporting concordance with conventional stereotactic and neurophysiologically guided targeting along the lateral axis in a small clinical cohort (Wu et al., 2026), underscoring growing clinical interest in ML-assisted stereotactic planning more broadly, albeit via a different (statistical-atlas rather than coordinate-regression) approach.

### 4.4 Model Generalizability and Comparisons

#### Validation for Other Imaging Modalities

By using paired datasets, we could directly compare predictions for different imaging modalities within the same participants. We found that prediction accuracy was maintained over a range of imaging modalities. These findings align with prior work from our group, which established the feasibility of accurately localizing AFIDs whether the MRI scans are research or clinical quality (Taha et al., 2023). Importantly, comparisons were conducted in AC-PC space, which assumes that independent AFID placements for paired modalities result in equivalent transforms. This approach was necessary due to the lack of ground truth STN segmentations in both the 1.5T MRI-gad and 3T MRI datasets. Despite this potential source of error, our results revealed no statistically significant differences in predicted coordinates.

#### Model Comparison and Performance Benchmarking

We employed a commonly used deformable registration pipeline to transform images from stereotactic space to native space. This pipeline employs rigorously optimized parameters (Ewert et al., 2019; Schönecker et al., 2009) for diffeomorphic registration from Advanced Normalization Tools (Avants et al., 2008), incorporating subcortical masks to refine deformation fields and enhance registration accuracy at (Ewert et al., 2019; Schönecker et al., 2009). Our model statistically outperformed registration when using 1.5T MRI-gad scans. MRI-gad scans can be considered a “lowest common denominator” scan employed for neurosurgical planning which provides anatomical detail and additionally the visualization of cerebral vasculature to ensure safe trajectory planning. However, the added gadolinium contrast introduces bias during registration (Abbass et al., 2022, 2025; Ogunsanya et al., 2024). Prior work comprehensively addressed the added benefit of multispectral imaging (Ewert et al., 2019). In this analysis, we employed sub-millimetric 7T T1w and T2w MRI data. Our model’s ability to match registration performance under these conditions highlights its potential utility in settings that lack access to 7T MRI. To our knowledge, our analysis represents the first millimetric account of registration error on MRI-gad scans at the STN. Future investigations should focus on a more comprehensive account of two elements pertaining to registration: (1) isolating and accounting for the MRI-gad bias introduced during registration by employing paired data and (2) evaluating which set of multispectral data provides the best advantage during registration.

#### Robustness to Annotation Variability

Not unexpectedly, our model performance was dependent on the accuracy of AFID localization. Indeed, model prediction error was found to increase linearly with AFID localization error (i.e., with every 1 mm of annotation error, prediction accuracy declines by approximately 0.46 mm; see Figure 4b). We believe the robustness to annotation error came from various factors: (1) PCA on input features, which attenuates the impact of isotropic noise by prioritizing structured anatomical variance and down-weighting noisy, low-variance directions; (2) redundancy among AFIDs, where overlapping spatial information allows the model to compensate for small errors in individual landmarks; and (3) regularization, which reduces model sensitivity to noisy features by constraining coefficient magnitudes.

#### Generalizability to Other Neuromodulation Targets

Beyond the STN, our framework maintained high millimetric localization accuracy for several additional subcortical targets, demonstrating the broad utility of AFID-based spatial relationships. This generalizability is valuable for targets that present significant visualization challenges with clinical imaging, such as the caudal zona incerta (cZI) and the fasciculus cerebellothalamicus (fct). The cZI cannot always be easily delineated from surrounding structures, surgical localization may rely on indirect, atlas-based targeting. Rather than targeting the cZI directly, neurosurgeons have used two neighboring structures that are highly visible on axial T2w scans (e.g., the red nucleus (RN) and subthalamic nucleus (STN)) as anatomical reference anchors to guide the placement of electrodes (Blomstedt et al., 2010; Plaha et al., 2006). These methods, however, present distinct limitations: both fail to account for inter-patient variability, and deformable atlas registration workflows can be computationally expensive and time consuming. Our coordinate-to-coordinate framework offers a fast and light-weight alternative to these indirect and atlas-based approaches (see Section 4.5 for the current validation status of this extension).

### 4.5 Limitations and Future Directions

A key limitation of our framework is its reliance on the accurate localization of AFIDs. However, prior studies have demonstrated that AFIDs can be placed reliably among a diverse rater demographics and MRI modalities. Furthermore, our detailed open-access placement protocol is designed to make AFID localization more accessible and standardized for a wider user base. Importantly, for our primary use case in DBS, neurosurgeons are accustomed to localization of AFIDs such as the AC and PC as part of routine clinical workflows. Extending these established practices to a broader set of landmarks within the AFID framework is therefore both feasible and well-aligned with current surgical expertise. Applying an automated detection step to input AFID landmarks is a natural next step and would enable an end-to-end pipeline from raw MRI to predicted surgical target. While the shared 16-AFID input is efficient and supported by the observed landmark redundancy (sections 3.4, 4.2), we have not benchmarked it against target-specific landmark subsets for cZI, fct, or RN; it remains possible that a differently optimized landmark set could further improve accuracy for these secondary targets, and the narrower validation scope applied to them relative to STN should be weighed accordingly when interpreting their reported accuracy. Unlike STN, where model accuracy was confirmed on a reserved test set separate from the cross-validation used for model selection, the reported cZI/fct/RN accuracy reflects the same cross-validation procedure used to select each identified winning architecture, with no independent held-out confirmation; these figures should be interpreted as an optimistic upper bound on true generalization accuracy for these targets rather than a confirmed estimate.

Although our model localizes targets with millimetric accuracy, it is important to recognize that optimal DBS outcomes are typically achieved by targeting the subregions of a given structure (e.g., dorsolateral STN in PD which corresponds to its motor functional territory). This spatial precision is crucial, as the true therapeutic substrate of DBS lies in modulating specific brain networks. In this work, we maintained an anatomically grounded framework as the STN center can be more objectively defined. However, our approach could be adapted to predict coordinates of sub-regions shown to have optimal therapeutic outcomes (Hollunder et al., 2024). Such extensions would allow the model to bridge anatomical and functional targeting strategies. We also emphasize that this framework predicts only the stereotactic target coordinate; it does not address trajectory or entry-point planning, which includes avoiding vessel or ventricle transgression as well as optimal DBS contact placement relative to the target.

Our current model is only geometry-based, relying on AFIDs without incorporating subject-level information that could enhance prediction accuracy or clinical relevance. Variables such as age, sex, diagnosis, disease severity, or even brain volume may further improve ML-driven localization accuracy. Future iterations of our framework could benefit from a hybrid approach, integrating demographic and clinical covariates alongside anatomical features to better tailor predictions to individual patients and potentially improve outcome prediction. Beyond the training-population and validation-scope limitations of the cZI/fct/RN extension already noted above, these models have also not been evaluated against prospective clinical outcomes, and the optimal active contact location may not coincide with any of these target centroids. Future iterations will expand the framework to include other common DBS targets to serve as a baseline benchmark alongside the fct and cZI models.

While DBS planning often involves implanting electrodes at predefined coordinates, this coordinate-based paradigm has limitations. In broader neuroscience contexts, many structures, particularly white matter pathways such as the fct, may not be fully captured by a single coordinate or point representation. Functional circuits, for instance, span or extend between anatomical locations, and their representation may be better suited to surface-based or tractography-informed models. Nevertheless, a coordinate-based representation of even a part of a white matter pathway can be considered a useful structural anchor in connectivity-based targeting. Future work may benefit from integrating volumetric, functional, or connectivity-based representations to more holistically capture this complexity.

## 5. Conclusions

In summary, we validated a coordinate-to-coordinate framework for the localization of deep brain structures including the STN and cZI. The model achieved millimetric precision, demonstrated strong generalizability on imaging modalities and additional subcortical landmarks, and outperformed conventional methods for brain structure localization on clinical scans. Its robustness to moderate annotation variability and low computational overhead make it well-suited for integration into neuroimaging and surgical planning pipelines. To facilitate accessibility and reproducibility, we have deployed the model as a user-friendly web application and openly released associated code and coordinate datasets.

## Supporting information

Supplementary Attachment C

## Data Availability

All data produced in the present study, trained models, analysis code, and scripts are openly available under the MIT License at https://github.com/afids/afids-pred, together with the annotation quality-control reports and an interactive 3D visualization of all coordinates.

## Data and Code Availability

All coordinate data, trained models, analysis code, and figure-generation scripts for this study, together with Supplementary Attachment A (HTML quality-control reports for the annotations; data/QC) and Supplementary Attachment B (interactive 3D visualization of all coordinates; figures/all_coordinates_interactive.html), are openly available under the MIT License at https://github.com/afids/afids-pred.

## Ethics Statement

Data collection was approved by the Western University Health Sciences Research Ethics Board (SNSX dataset, REB# 109045) and the London Health Sciences Centre Research Ethics Board (LHSCPD dataset, REB# R-17-156), as described in Section 2.1. All participants provided written informed consent.

## Author Contributions (CRediT)

Conceptualization: AT, ARK, JCL. Data curation: AT, JT, MA, JZ, EC, VML, ArT, GG Formal analysis: AT, JT. Funding acquisition: AT, ARK, JCL. Investigation: AT, JT, MA, ArT, MS, VML. Methodology: AT, JT, ARK, JCL. Project administration: JCL. Resources: MA, GG, ARK, JCL. Software: AT, JT, MS, DB, GG. Supervision: ARK, JCL. Validation: AT, JZ, EC. Visualization: AT. Writing – original draft: AT. Writing – review & editing: all authors.

## Acknowledgements

The authors thank the raters who contributed landmark annotations and the participants who took part in this research. The authors also acknowledge the imaging support provided by the Centre for Functional and Metabolic Mapping (CFMM), Robarts Research Institute, Western University.

## Supplementary Material

Supplementary Methods S1–S2, Supplementary Results S3, Supplementary Figures S1–S6, and Supplementary Tables S1–S5 are provided in a separate Supplementary Material file. Supplementary Attachment A (quality-control reports) and Supplementary Attachment B (interactive 3D visualization of all coordinates) are available online (https://github.com/afids/afids-pred; see Data and Code Availability), and Supplementary Attachment C (16-AFID placement protocol) is provided as a separate file.

## References

Abbass, M., Gilmore, G., Taha, A., Chevalier, R., Jach, M., Peters, T. M., Khan, A. R., & Lau, J. C. (2022). Application of the anatomical fiducials framework to a clinical dataset of patients with Parkinson’s disease. Brain Structure & Function, 227(1), 393–405. 10.1007/s00429-021-02408-3

Abbass, M., Taha, A., Gilmore, G., Santyr, B., Chalil, A., Jog, M., MacDougall, K., Parrent, A. G., Peters, T. M., & Lau, J. C. (2025). The impact of localization and registration accuracy on estimates of deep brain stimulation electrode position in stereotactic space. Imaging Neuroscience, 3, Article imag_a_00579. 10.1162/imag_a_00579

Abosch, A., Yacoub, E., Ugurbil, K., & Harel, N. (2010). An assessment of current brain targets for deep brain stimulation surgery with susceptibility-weighted imaging at 7 tesla. Neurosurgery, 67(6), 1745–1756; discussion 1756. 10.1227/neu.0b013e3181f74105

Alkemade, A., de Hollander, G., Keuken, M. C., Schäfer, A., Ott, D. V. M., Schwarz, J., Weise, D., Kotz, S. A., & Forstmann, B. U. (2017). Comparison of T2*-weighted and QSM contrasts in Parkinson’s disease to visualize the STN with MRI. PLOS ONE, 12(4), e0176130. 10.1371/journal.pone.0176130

Anheim, M., Batir, A., Fraix, V., Silem, M., Chabardès, S., Seigneuret, E., Krack, P., Benabid, A.-L., & Pollak, P. (2008). Improvement in Parkinson disease by subthalamic nucleus stimulation based on electrode placement: Effects of reimplantation. Archives of Neurology, 65(5), 612–616. 10.1001/archneur.65.5.612

Avants, B. B., Epstein, C. L., Grossman, M., & Gee, J. C. (2008). Symmetric diffeomorphic image registration with cross-correlation: evaluating automated labeling of elderly and neurodegenerative brain. Medical Image Analysis, 12(1), 26–41. 10.1016/j.media.2007.06.004

Baniasadi, M., Petersen, M. V., Gonçalves, J., Horn, A., Vlasov, V., Hertel, F., & Husch, A. (2023). DBSegment: Fast and robust segmentation of deep brain structures considering domain generalization. Human Brain Mapping, 44(2), 762–778. 10.1002/hbm.26097

Blomstedt, P., Sandvik, U., & Tisch, S. (2010). Deep brain stimulation in the posterior subthalamic area in the treatment of essential tremor. Movement Disorders, 25(10), 1350–1356. 10.1002/mds.22758

Boutet, A., Loh, A., Chow, C. T., Taha, A., Elias, G. J. B., Neudorfer, C., Germann, J., Paff, M., Zrinzo, L., Fasano, A., Kalia, S. K., Steele, C. J., Mikulis, D., Kucharczyk, W., & Lozano, A. M. (2021). A literature review of magnetic resonance imaging sequence advancements in visualizing functional neurosurgery targets. Journal of Neurosurgery, 135(5), 1445–1458. 10.3171/2020.8.jns201125

Buhmann, C., Huckhagel, T., Engel, K., Gulberti, A., Hidding, U., Poetter-Nerger, M., Goerendt, I., Ludewig, P., Braass, H., Choe, C.-U., Krajewski, K., Oehlwein, C., Mittmann, K., Engel, A. K., Gerloff, C., Westphal, M., Köppen, J. A., Moll, C. K. E., & Hamel, W. (2017). Adverse events in deep brain stimulation: A retrospective long-term analysis of neurological, psychiatric and other occurrences. PLOS ONE, 12(7), e0178984. 10.1371/journal.pone.0178984

Camlidag, I., Kocabicak, E., Sahin, B., Jahanshahi, A., Incesu, L., Aygun, D., Yildiz, O., Temel, Y., & Belet, U. (2014). Volumetric analysis of the subthalamic and red nuclei based on magnetic resonance imaging in patients with Parkinson’s disease. The International Journal of Neuroscience, 124(4), 291–295. 10.3109/00207454.2013.843091

Chakravarty, M. M., Bertrand, G., Hodge, C. P., Sadikot, A. F., & Collins, D. L. (2006). The creation of a brain atlas for image guided neurosurgery using serial histological data. NeuroImage, 30(2), 359–376. 10.1016/j.neuroimage.2005.09.041

Chandran, A. S., Bynevelt, M., & Lind, C. R. P. (2016). Magnetic resonance imaging of the subthalamic nucleus for deep brain stimulation. Journal of Neurosurgery, 124(1), 96–105. 10.3171/2015.1.jns142066

Chen, T., & Guestrin, C. (2016). XGBoost: A scalable tree boosting system. In Proceedings of the 22nd ACM SIGKDD International Conference on Knowledge Discovery and Data Mining (pp. 785–794). Association for Computing Machinery. 10.1145/2939672.2939785

Chen, X., Qu, L., Xie, Y., Ahmad, S., & Yap, P.-T. (2023). A paired dataset of T1- and T2-weighted MRI at 3 Tesla and 7 Tesla. Scientific Data, 10(1), 489. 10.1038/s41597-023-02400-y

Clarke, W. T., Mougin, O., Driver, I. D., Rua, C., Morgan, A. T., Asghar, M., Clare, S., Francis, S., Wise, R. G., Rodgers, C. T., Carpenter, A., Muir, K., & Bowtell, R. (2020). Multi-site harmonization of 7 Tesla MRI neuroimaging protocols. NeuroImage, 206, 116335. 10.1016/j.neuroimage.2019.116335

Collins, D. L., Neelin, P., Peters, T. M., & Evans, A. C. (1994). Automatic 3-D intersubject registration of MR volumetric data in standardized Talairach space. Journal of Computer Assisted Tomography, 18(2), 192–205. 10.1097/00004728-199403000-00005

Colpan, M. E., & Slavin, K. V. (2010). Subthalamic and red nucleus volumes in patients with Parkinson’s disease: do they change with disease progression? Parkinsonism & Related Disorders, 16(6), 398–403. 10.1016/j.parkreldis.2010.03.008

Duchin, Y., Abosch, A., Yacoub, E., Sapiro, G., & Harel, N. (2012). Feasibility of using ultra-high field (7 T) MRI for clinical surgical targeting. PLOS ONE, 7(5), e37328. 10.1371/journal.pone.0037328

Duchin, Y., Shamir, R. R., Patriat, R., Kim, J., Vitek, J. L., Sapiro, G., & Harel, N. (2018). Patient-specific anatomical model for deep brain stimulation based on 7 Tesla MRI. PLOS ONE, 13(8), e0201469. 10.1371/journal.pone.0201469

Edlow, B. L., Mareyam, A., Horn, A., Polimeni, J. R., Witzel, T., Tisdall, M. D., Augustinack, J. C., Stockmann, J. P., Diamond, B. R., Stevens, A., Tirrell, L. S., Folkerth, R. D., Wald, L. L., Fischl, B., & van der Kouwe, A. (2019). 7 Tesla MRI of the ex vivo human brain at 100 micron resolution. Scientific Data, 6(1), 244. 10.1038/s41597-019-0254-8

Engelhardt, J., Cuny, E., Guehl, D., Burbaud, P., Damon-Perrière, N., Dallies-Labourdette, C., Thomas, J., Branchard, O., Schmitt, L.-A., Gassa, N., & Zemzemi, N. (2021). Prediction of Clinical Deep Brain Stimulation Target for Essential Tremor From 1.5 Tesla MRI Anatomical Landmarks. Frontiers in Neurology, 12, 620360. 10.3389/fneur.2021.620360

Ewert, S., Horn, A., Finkel, F., Li, N., Kühn, A. A., & Herrington, T. M. (2019). Optimization and comparative evaluation of nonlinear deformation algorithms for atlas-based segmentation of DBS target nuclei. NeuroImage, 184, 586–598. 10.1016/j.neuroimage.2018.09.061

Ewert, S., Plettig, P., Li, N., Chakravarty, M. M., Collins, D. L., Herrington, T. M., Kühn, A. A., & Horn, A. (2018). Toward defining deep brain stimulation targets in MNI space: A subcortical atlas based on multimodal MRI, histology and structural connectivity. NeuroImage, 170, 271–282. 10.1016/j.neuroimage.2017.05.015

Fedorov, A., Beichel, R., Kalpathy-Cramer, J., Finet, J., Fillion-Robin, J.-C., Pujol, S., Bauer, C., Jennings, D., Fennessy, F., Sonka, M., Buatti, J., Aylward, S., Miller, J. V., Pieper, S., & Kikinis, R. (2012). 3D Slicer as an image computing platform for the Quantitative Imaging Network. Magnetic Resonance Imaging, 30(9), 1323–1341. 10.1016/j.mri.2012.05.001

Fonov, V. S., Evans, A. C., McKinstry, R. C., Almli, C. R., & Collins, D. L. (2009). Unbiased nonlinear average age-appropriate brain templates from birth to adulthood. NeuroImage, 47(Suppl. 1), S102. 10.1016/s1053-8119(09)70884-5

Friedman, J. H. (2001). Greedy function approximation: A gradient boosting machine. Annals of Statistics, 29(5), 1189–1232. 10.1214/aos/1013203451

Guo, T., Parrent, A. G., & Peters, T. M. (2007). Surgical targeting accuracy analysis of six methods for subthalamic nucleus deep brain stimulation. Computer Aided Surgery: Official Journal of the International Society for Computer Aided Surgery, 12(6), 325–334. 10.3109/10929080701730987

Hoerl, A. E., & Kennard, R. W. (1970). Ridge regression: Biased estimation for nonorthogonal problems. Technometrics: A Journal of Statistics for the Physical, Chemical, and Engineering Sciences, 12(1), 55–67. 10.1080/00401706.1970.10488634

Hollunder, B., Ostrem, J. L., Sahin, I. A., Rajamani, N., Oxenford, S., Butenko, K., Neudorfer, C., Reinhardt, P., Zvarova, P., Polosan, M., Akram, H., Vissani, M., Zhang, C., Sun, B., Navratil, P., Reich, M. M., Volkmann, J., Yeh, F.-C., Baldermann, J. C., … Horn, A. (2024). Mapping dysfunctional circuits in the frontal cortex using deep brain stimulation. Nature Neuroscience, 27(3), 573–586. 10.1038/s41593-024-01570-1

Horsley, V., & Clarke, R. H. (1908). The structure and functions of the cerebellum examined by a new method. Brain: A Journal of Neurology, 31(1), 45–124. 10.1093/brain/31.1.45

Kaya, M. O., Ozturk, S., Ercan, I., Gonen, M., Serhat Erol, F., & Kocabicak, E. (2019). Statistical shape analysis of subthalamic nucleus in patients with Parkinson disease. World Neurosurgery, 126, e835–e841. 10.1016/j.wneu.2019.02.180

Kremer, N. I., van Laar, T., Lange, S. F., Statius Muller, S., la Bastide-van Gemert, S., Oterdoom, D. L. M., Drost, G., & van Dijk, J. M. C. (2023). STN-DBS electrode placement accuracy and motor improvement in Parkinson’s disease: systematic review and individual patient meta-analysis. Journal of Neurology, Neurosurgery, and Psychiatry, 94(3), 236–244. 10.1136/jnnp-2022-329192

Lau, J. C., Khan, A. R., Zeng, T. Y., MacDougall, K. W., Parrent, A. G., & Peters, T. M. (2018). Quantification of local geometric distortion in structural magnetic resonance images: Application to ultra-high fields. NeuroImage, 168, 141–151. 10.1016/j.neuroimage.2016.12.066

Lau, J. C., MacDougall, K. W., Arango, M. F., Peters, T. M., Parrent, A. G., & Khan, A. R. (2017). Ultra-High Field Template-Assisted Target Selection for Deep Brain Stimulation Surgery. World Neurosurgery, 103, 531–537. 10.1016/j.wneu.2017.04.043

Lau, J. C., Parrent, A. G., Demarco, J., Gupta, G., Kai, J., Stanley, O. W., Kuehn, T., Park, P. J., Ferko, K., Khan, A. R., & Peters, T. M. (2019). A framework for evaluating correspondence between brain images using anatomical fiducials. Human Brain Mapping, 40(14), 4163–4179. 10.1002/hbm.24693

Lau, J. C., Xiao, Y., Haast, R. A. M., Gilmore, G., Uludağ, K., MacDougall, K. W., Menon, R. S., Parrent, A. G., Peters, T. M., & Khan, A. R. (2020). Direct visualization and characterization of the human zona incerta and surrounding structures. Human Brain Mapping, 41(16), 4500–4517. 10.1002/hbm.25137

Lenglet, C., Abosch, A., Yacoub, E., De Martino, F., Sapiro, G., & Harel, N. (2012). Comprehensive in vivo mapping of the human basal ganglia and thalamic connectome in individuals using 7T MRI. PLOS ONE, 7(1), e29153. 10.1371/journal.pone.0029153

Lozano, A. M., Lipsman, N., Bergman, H., Brown, P., Chabardes, S., Chang, J. W., Matthews, K., McIntyre, C. C., Schlaepfer, T. E., Schulder, M., Temel, Y., Volkmann, J., & Krauss, J. K. (2019). Deep brain stimulation: current challenges and future directions. Nature Reviews Neurology, 15(3), 148–160. 10.1038/s41582-018-0128-2

Middlebrooks, E. H., Bot, M., Patriat, R., Lau, J. C., Grewal, S. S., Zhou, X., Tao, S., Straub, S., Yacoub, E., McGovern, R., & Harel, N. (2025). Multi-institutional recommendations on the use of 7T MRI in deep brain stimulation. Journal of Neurosurgery, 143(5), 1165–1175. 10.3171/2025.3.jns243024

Miller, C. P. K., Muller, J., Noecker, A. M., Matias, C., Alizadeh, M., McIntyre, C., & Wu, C. (2023). Automatic segmentation of Parkinson disease therapeutic targets using nonlinear registration and clinical MR imaging: Comparison of methodology, presence of disease, and quality control. Stereotactic and Functional Neurosurgery, 101(2), 146–157. 10.1159/000526719

Neudorfer, C., Butenko, K., Oxenford, S., Rajamani, N., Achtzehn, J., Goede, L., Hollunder, B., Ríos, A. S., Hart, L., Tasserie, J., Fernando, K. B., Nguyen, T. A. K., Al-Fatly, B., Vissani, M., Fox, M., Richardson, R. M., van Rienen, U., Kühn, A. A., Husch, A. D., … Horn, A. (2023). Lead-DBS v3.0: Mapping deep brain stimulation effects to local anatomy and global networks. NeuroImage, 268, 119862. 10.1016/j.neuroimage.2023.119862

Ogunsanya, F., Taha, A., Gilmore, G., Kai, J., Kuehn, T., Thurairajah, A., Tenorio, M. C., Khan, A. R., & Lau, J. C. (2024). MRI-degad: toward accurate conversion of gadolinium-enhanced T1w MRIs to non-contrast-enhanced scans using CNNs. International Journal of Computer Assisted Radiology and Surgery, 19(7), 1469–1472. 10.1007/s11548-024-03186-z

Párraga, R. G., Possatti, L. L., Alves, R. V., Ribas, G. C., Türe, U., & de Oliveira, E. (2016). Microsurgical anatomy and internal architecture of the brainstem in 3D images: surgical considerations. Journal of Neurosurgery, 124(5), 1377–1395. 10.3171/2015.4.jns132778

Patriat, R., Niederer, J., Kaplan, J., Amundsen Huffmaster, S., Petrucci, M., Eberly, L., Harel, N., & MacKinnon, C. (2020). Morphological changes in the subthalamic nucleus of people with mild-to-moderate Parkinson’s disease: a 7-T MRI study. Scientific Reports, 10(1), 8785. 10.1038/s41598-020-65752-0

Pedregosa, F., Varoquaux, G., Gramfort, A., Michel, V., Thirion, B., Grisel, O., Blondel, M., Prettenhofer, P., Weiss, R., Dubourg, V., Vanderplas, J., Passos, A., Cournapeau, D., Brucher, M., Perrot, M., & Duchesnay, É. (2011). Scikit-learn: Machine learning in Python. Journal of Machine Learning Research, 12, 2825–2830.

Perera, I. R., Zahed, M., Moriarty, S., Simmons, Z., Rodriguez, M., Botkin, C., Dickson, T., Kasper, B., Fahmy, K., & Millard, J. A. (2024). Geometric morphometric analysis of the brainstem and cerebellum in Chiari I malformation. Frontiers in Neuroanatomy, 18, 1434017. 10.3389/fnana.2024.1434017

Plaha, P., Ben-Shlomo, Y., Patel, N. K., & Gill, S. S. (2006). Stimulation of the caudal zona incerta is superior to stimulation of the subthalamic nucleus in improving contralateral parkinsonism. Brain, 129(7), 1732–1747. 10.1093/brain/awl127

Reich, M. M., Hsu, J., Ferguson, M., Schaper, F. L. W. V. J., Joutsa, J., Roothans, J., Nickl, R. C., Frankemolle-Gilbert, A., Alberts, J., Volkmann, J., & Fox, M. D. (2022). A brain network for deep brain stimulation induced cognitive decline in Parkinson’s disease. Brain: A Journal of Neurology, 145(4), 1410–1421. 10.1093/brain/awac012

Ren, J., An, N., Lin, C., Zhang, Y., Sun, Z., Zhang, W., Li, S., Guo, N., Cui, W., Hu, Q., Wang, W., Wu, X., Wang, Y., Jiang, T., Satterthwaite, T. D., Wang, D., & Liu, H. (2025). DeepPrep: an accelerated, scalable and robust pipeline for neuroimaging preprocessing empowered by deep learning. Nature Methods, 22(3), 473–476. 10.1038/s41592-025-02599-1

Schaltenbrand, G., Wahren, W., & Hassler, R. G. (1977). Atlas for stereotaxy of the human brain. Thieme.

Schönecker, T., Kupsch, A., Kühn, A. A., Schneider, G.-H., & Hoffmann, K.-T. (2009). Automated optimization of subcortical cerebral MR imaging-atlas coregistration for improved postoperative electrode localization in deep brain stimulation. AJNR. American Journal of Neuroradiology, 30(10), 1914–1921. 10.3174/ajnr.a1741

Taha, A., Gilmore, G., Abbass, M., Kai, J., Kuehn, T., Demarco, J., Gupta, G., Zajner, C., Cao, D., Chevalier, R., Ahmed, A., Hadi, A., Karat, B. G., Stanley, O. W., Park, P. J., Ferko, K. M., Hemachandra, D., Vassallo, R., Jach, M., … Lau, J. C. (2023). Magnetic resonance imaging datasets with anatomical fiducials for quality control and registration. Scientific Data, 10(1), 449. 10.1038/s41597-023-02330-9

Talairach, J., David, M., Tournoux, P., Corredor, H., & Kvasina, T. (1957). Atlas d’anatomie stéréotaxique: Repérage radiologique indirect des noyaux gris centraux des régions mésencéphalo-sous-optique et hypothalamique de l’homme. Masson.

Warntjes, J. B. M., Tisell, A., Landtblom, A.-M., & Lundberg, P. (2014). Effects of gadolinium contrast agent administration on automatic brain tissue classification of patients with multiple sclerosis. AJNR. American Journal of Neuroradiology, 35(7), 1330–1336. 10.3174/ajnr.a3890

Wu, F., Buonanno, R., Baro, V., Levi, V., Furlanis, G. M., Gagliano, M., Guerra, A., D’Amico, A., Giussani, C. G., Eleopra, R., Denaro, L., Antonini, A., & Landi, A. (2026). Artificial intelligence for STN-DBS surgical planning in Parkinson’s disease: A multicenter study comparing conventional targeting versus supervised statistical machine learning. Brain Sciences, 16(5), 457. 10.3390/brainsci16050457

