## Supplementary Material for "A coordinate-to-coordinate machine learning framework for contrast-agnostic brain target localization"

All in-text citations refer to the reference list of the main manuscript. Analysis code for every result in this document is in the afids-pred repository https://github.com/afids/afids-pred; notebooks/supplementary_analyses.py; notebooks/2.6_Generalized_Target_Prediction_unified.py).

#### Contents

- Supplementary Methods S1. Machine learning methodology for additional proof-of-concept targets (cZI, fct, RN)
- Supplementary Methods S2. Supplementary analyses of the STN model
- Supplementary Results S3
- Supplementary Tables S1–S5
- Supplementary Figures S1–S6
- **Supplementary Attachments A–B** (online, https://github.com/afids/afids-pred) and C (separate file)

#### Supplementary Methods S1. Machine learning methodology for additional proof-of-concept targets

##### S1.1 Datasets

Model training and development used the healthy controls subset of the Stereotactic Neurosurgery (SNSX) dataset, consisting of 32 Ultra-High field strength (7 Tesla) T1-weighted scans (mean age: 46.2 ± 13.5 years; median: 48 years; range: 20–70 years; 12 female and 20 male; right-handed). Ground truth target output coordinates (x,y,z) were extracted from manually curated 7-T segmentations for each of the following targets: cZI, fct, and RN. Unlike the primary STN model (main text, section 2.1), which includes both healthy control and Parkinson's disease participants, training and evaluation for these secondary targets was restricted to the healthy-control subset because manually curated cZI, fct, and RN segmentations were only available for these participants; this should be considered when interpreting the generalizability of these models to clinical (e.g., disease-affected) populations, since STN accuracy was validated on a clinically representative cohort while cZI/fct/RN accuracy has not yet been.

##### S1.2 Feature selection and data preprocessing

This extension uses the same manually curated 16-AFID subset as the primary STN model described in main-text section 2.2, ensuring methodological consistency across all coordinate-to-coordinate targets in this framework (Supplementary Figure S1).

All training coordinate data underwent AC-PC alignment and mid-commissural point (MCP) centering transformations. The coordinates on the left hemisphere were flipped to the right by inverting the sign of the X-coordinate after alignment, providing two training examples per subject (64 training instances across 32 participants for cZI and fct; 62 across 31 participants for RN). As with the primary STN model (main-text section 2.4), a single shared StandardScaler-then-PCA (99%-variance) pipeline, refit within each training fold on the fold's training examples plus an AFID-only pool of all AFID scans outside the target cohort, was applied uniformly across all ten candidate models during the model-comparison screen (see section S1.3.1).

#### S1.3 Training and cross-validation

##### S1.3.1 Model suite

Three separate investigations were conducted, one for the cZI, one for the RN, and one for the fct. Each investigation evaluated ten regression models to determine the optimal mapping function between the 16-AFID subset (the same input features used by the primary STN model) and their respective ground-truth target coordinates (x,y,z). For each of these ten models, separate and independent regression models were trained for each spatial dimension (x,y, and z). As with the STN model comparison (main-text section 2.4), a single shared preprocessing pipeline (StandardScaler followed by PCA retaining 99% of variance, refit within each training fold) was applied uniformly across all ten candidate models.

The candidate models evaluated were: Ridge, BayesianRidge, ElasticNet, RandomForest, Lasso, Partial Least Squares (PLS), XGBoost, Support Vector Regression with RBF Kernel (SVR), K-Nearest Neighbors (KNN) and Multi-Layer Perceptron (MLP).

Hyperparameters were tuned inside each outer training fold with 5-fold participant-grouped inner cross-validation. The search spaces were: Ridge, $\alpha \in 10^{-3}$–$10^{6}$ (200 log-spaced values); Lasso, $\alpha \in 10^{-3}$–$10^{1}$ (50 values); ElasticNet, $\alpha \in 10^{-3}$–$10^{1}$ (30 values) × l1-ratio ∈ {0.1–1.0}; PLS, 1–10 components; KNN, k = 1–10 with uniform or distance weights; RandomForest, XGBoost, SVR and MLP, randomized search (12, 20, 20 and 10 iterations). BayesianRidge estimates its regularization by evidence maximization and was fitted without a search.

##### S1.3.2 Leave-One-Group-Out Cross-Validation (LOGO-CV)

Models were evaluated using a Leave-One-Group-Out Cross-validation scheme grouped by subject ID (32 folds for cZI and fct; 31 folds for RN). Grouping by subject ensured that both hemispheric training examples were kept together in either the training set or the validation set for each fold, to prevent any data leakage. The models were evaluated by computing the ED between the predicted target coordinates and their ground-truth centroid coordinates in native space. (For RN, one of the 32 training subjects lacked a usable ground-truth annotation and was excluded, giving 31 participants, 31 folds and 62 hemisphere estimates for that target; see main-text sections 2.6 and 3.5.) As with the STN model comparison (Supplementary Table S2), per-fold results were retained and pairwise Holm-Bonferroni-corrected Wilcoxon signed-rank tests against each target's winning model are reported in Supplementary Tables S3–S5 and Supplementary Figure S2.

The candidates were closely clustered, and the ranking depends on the summary statistic. Ranked by mean rather than median ED, Ridge would be selected for fct (mean 0.798 vs. 0.887 mm for RandomForest) and for RN (mean 0.515 vs. 0.561 mm for Lasso). The selected models should therefore be read as the best point estimates among near-equivalent candidates, not as statistically confirmed winners. Because model selection and accuracy estimation used the same cross-validation, the reported accuracies are optimistic.

#### S1.4 Model deployment

The top-performing models are ElasticNet for cZI, RandomForest for fct, and Lasso for RN and were retrained on the full training dataset (64 instances across 32 subjects for cZI and fct; 62 instances across 31 subjects for RN). The finalized models were packaged and exported as .pkl files for deployment. The web tool accepts AFIDs placed in native image space and derives the AC-PC/MCP transformation itself. At the time of writing, the stereotaxy.afids.io web tool returns STN predictions only; the cZI, fct and RN models are distributed as .pkl files in the afids-pred repository and have not yet been integrated into the web interface.

### Supplementary Methods S2. Supplementary analyses of the STN model

#### S2.1 Common setup

All analyses use the production pipeline (StandardScaler → PCA retaining 99% of variance → Ridge regression per axis, with α selected by 3-fold participant-grouped inner cross-validation over 200 log-spaced values in $10^{-3}$–$10^{6}$). The deployed model was tuned with an earlier version of the code over a wider grid ($10^{-100}$–$10^{100}$); its selected α values, 4.6–11.6, lie inside both ranges. The training set comprised 50 SNSX participants (32 healthy controls, 18 Parkinson's disease; 100 hemisphere examples). PCA was fit on the training examples plus 224 AFID-only hemisphere examples from scans outside both the training and test sets. Cross-validation folds were always grouped by participant. Held-out evaluations used the 80 test STN localizations of main-text Table 2 (four acquisitions × 10 participants × 2 hemispheres), with 1.5-T gadolinium-enhanced and 3-T predictions scored against the paired 7-T STN annotations.

**Note on grouping.** An earlier version of the STN model screen, of the Ridge–XGBoost comparison and of the leave-one-AFID-out analysis assigned cross-validation groups by row position rather than by participant identity, which placed the two (mirrored) hemisphere examples of some participants in different folds. All results in this document were recomputed with participant-level grouping and supersede those values.

#### S2.2 Architecture screen and Ridge–XGBoost comparison

The ten-model screen used the roster and per-model hyperparameter searches of S1.3, with 3-fold participant-grouped inner cross-validation. The outer loop was 4-fold participant-grouped cross-validation repeated five times with different random assignments of participants to folds. Each hemisphere's ED was averaged over the five repeats. Because the two hemispheres of a participant are not independent, models were compared with paired Wilcoxon signed-rank tests on participant-mean ED ($n = 50$), Holm-Bonferroni corrected against Ridge (Supplementary Table S2; Supplementary Figure S2). Ridge and XGBoost were additionally compared on per-axis squared error and ED with the same participant-level pairing, Bonferroni corrected across the four metrics (Supplementary Figure S3). Out-of-fold cross-validation errors were also compared between healthy-control and Parkinson's disease participants (Mann-Whitney U test on participant-mean ED).

#### S2.3 Pipeline sensitivity

**PCA threshold.** The pipeline was refit with PCA retaining 80%, 90%, 95%, 99%, 99.5% or 99.9% of variance, or without PCA, and evaluated by training cross-validation (5 × 4-fold grouped) and on the held-out test set. All sensitivity analyses refit the pipeline from the training data with the current code; the refit production configuration therefore differs slightly from the deployed model on the held-out set (median ED 1.22 vs. 1.20 mm), which was fitted with an earlier version of the code.

**Learning curve.** Training sets of 8–50 participants were drawn at random without replacement (25 draws per size; one at $n = 50$), each used to fit the full pipeline, and each fit was scored on the fixed held-out test set.

#### S2.4 Landmark importance

Ablation importance was the change in cross-validated median ED (4-fold, participant-grouped) when all three coordinates of one of the 11 encoded landmarks were removed and the pipeline refit. To estimate the accuracy of a reduced landmark set without selection bias, greedy forward selection was nested inside cross-validation. Within each outer training fold, landmarks were added one at a time according to 3-fold inner-CV median ED. After each addition, the current subset was refit on the outer training fold and scored on the untouched outer test fold.

#### S2.5 Leave-one-AFID-out landmark reconstruction

To test whether the coordinate-to-coordinate approach generalizes beyond the STN (main text, sections 2.5 and 3.4), each of the 11 encoded landmarks was in turn treated as the prediction target (x, y, z) and predicted from the coordinates of the remaining ten with the production pipeline (PCA fit on the training examples plus the AFID-only pool, restricted to the predictor coordinates). The analysis used the 100 hemisphere examples of the 50 training participants and 5 × 4-fold cross-validation grouped by participant identity, with each example's ED averaged over the five repeats. As a reference, each landmark was also 'predicted' by its mean position in the training fold. LVAC and LVPC lie on the lateral ventricles rather than the midbrain; as in the original analysis they are shown but excluded from the pooled midbrain summary. In AC–PC/MCP-centered space, AC and PC have $x = z = 0$ and $ACy = -PCy$ by construction, so each is an exact linear function of the other and their reconstruction is trivial; PMJx is likewise fixed at 0 by the midsagittal alignment.

##### S2.6 Precision of the held-out estimate

The 80 test localizations come from 20 participants, each contributing two hemispheres and two acquisitions. A 95% confidence interval for the test-set median ED was therefore obtained by a participant-level cluster bootstrap (10,000 resamples of the 20 participants, retaining all of a participant's localizations).

#### Supplementary Results S3

**S3.1 Held-out accuracy.** Across the 80 test STNs the median ED was 1.20 mm (SD 0.44; IQR 0.84–1.53 mm; participant-level bootstrap 95% CI 1.01–1.38 mm), and 98.8% of localizations were within 2 mm.

**S3.2 Architecture screen.** Ridge had the lowest median cross-validated ED (0.85 mm; Supplementary Table S2; Supplementary Figure S2). After Holm correction it was not distinguishable from BayesianRidge, PLS, RandomForest, or ElasticNet, and it was more accurate than SVR, Lasso, XGBoost, KNN, and MLP. Against XGBoost, Ridge had lower ED ($p = 0.002$) and lower squared error along x ($p = 0.011$) and y ($p = 0.006$), with no difference along z ($p = 1.00$; Bonferroni-corrected; Supplementary Figure S3).

**S3.3 Health status.** Cross-validated error was higher in Parkinson's disease than in healthy-control training participants (median 1.13 vs. 0.80 mm; Mann-Whitney $p = 0.003$). This is consistent with the held-out test set, which consists entirely of Parkinson's disease patients, having higher error than training cross-validation.

**S3.4 Pipeline sensitivity.** Test-set median ED varied between 1.22 and 1.29 mm across PCA thresholds from 80% to 99.9% and without PCA (Supplementary Figure S5). The 99% threshold (24 components) had the lowest training cross-validation error. In the learning curve, median test ED fell from 1.48 mm with 8 training participants to 1.29 mm at 30 and 1.22 mm at 50.

**S3.5 Landmark importance.** Removing LVPC, PMJ, LVAC, SLMS, SIPF or MB increased cross-validated error (+0.071 mm, +0.049 mm, +0.043 mm, +0.041 mm, +0.015 mm, +0.011 mm), whereas removing any other single landmark changed it by ≤ 0.019 mm, reflecting the redundancy among AFIDs (main text, section 4.2; Supplementary Figure S6). In nested cross-validation the lowest outer-fold median ED was 0.85 mm with 4 landmarks, against 0.88 mm with all 11. Landmarks most often chosen among the first five were SLMS (4/4 folds), LVPC (3/4 folds), MB (3/4 folds), and AC (2/4 folds).

**S3.6 Leave-one-AFID-out landmark reconstruction.** Pooled across the nine midbrain landmarks, the median reconstruction error was 0.80 mm (IQR 0.34–1.33 mm; mean 1.02 ± 1.48 mm; Supplementary Figure S4). AC and PC were reconstructed almost exactly (median 0.063 and 0.063 mm), which reflects the AC–PC frame rather than learned anatomy (S2.5). Excluding them, the pooled median over the remaining seven midbrain landmarks was 1.00 mm (IQR 0.67–1.53 mm). By landmark, MB 0.67 mm, IMS 0.68 mm, SIPF 0.79 mm, ICS 1.17 mm, SLMS 1.28 mm, ILMS 1.38 mm, PMJ 1.64 mm (median ED, in increasing order). The ventricular landmarks were reconstructed less accurately (LVAC 1.92 mm; LVPC 2.01 mm).

#### Supplementary Tables

**Supplementary Table S1.** Summary of anatomical fiducial (AFID) and subthalamic nucleus (STN) annotation across datasets. Each row represents a unique annotator (Rater ID) contributing to one or more datasets. The table includes the experience (in months) of raters in three domains: Imaging, Neuroanatomy, and 3D Slicer. Rater ID entries are context-specific and assigned uniquely within each dataset.

| Annotation | Dataset | Rater ID | Imaging (mo) | Neuroanatomy (mo) | 3D Slicer (mo) |
|---|---|---|---|---|---|
| AFIDs | LHSCPD | AT | 0 | 0 | 0 |
| AFIDs | LHSCPD | GG | 60 | 60 | 60 |
| AFIDs | LHSCPD | MA | 60 | 60 | 60 |
| AFIDs | LHSCPD | MJ | 0 | 0 | 0 |
| AFIDs | LHSCPD | RC | 0 | 0 | 0 |
| AFIDs | AFIDs-HCP | AT | 60 | 60 | 60 |
| AFIDs | AFIDs-HCP | CZ | 12 | 12 | 12 |
| AFIDs | AFIDs-HCP | GG | 60 | 60 | 60 |
| AFIDs | AFIDs-HCP | DC | 24 | 24 | 24 |
| AFIDs | AFIDs-HCP | KF | 24 | 24 | 24 |
| AFIDs | SNSX (healthy controls) | Rater01 | 12 | 12 | 12 |
| AFIDs | SNSX (healthy controls) | Rater02 | 24 | 24 | 24 |
| AFIDs | SNSX (healthy controls) | Rater03 | 24 | 24 | 24 |
| AFIDs | SNSX (healthy controls) | Rater04 | 36 | 36 | 36 |
| AFIDs | SNSX (healthy controls) | Rater05 | 24 | 24 | 24 |
| AFIDs | SNSX (healthy controls) | Rater06 | 24 | 24 | 24 |
| AFIDs | SNSX (healthy controls) | Rater07 | 24 | 24 | 24 |
| AFIDs | SNSX (healthy controls) | Rater08 | 60 | 60 | 60 |
| AFIDs | SNSX (healthy controls) | Rater09 | 12 | 12 | 12 |
| AFIDs | AFIDs-OASIS, MNI | Rater01 | 24 | 24 | 24 |
| AFIDs | AFIDs-OASIS, MNI | Rater02 | 0 | 0 | 0 |
| AFIDs | AFIDs-OASIS, MNI | Rater03 | 8 | 0 | 8 |
| AFIDs | AFIDs-OASIS, MNI | Rater04 | 24 | 6 | 0 |
| AFIDs | AFIDs-OASIS, MNI | Rater05 | 0 | 24 | 0 |
| AFIDs | AFIDs-OASIS, MNI | Rater06 | 24 | 12 | 12 |
| AFIDs | AFIDs-OASIS, MNI | Rater07 | 12 | 48 | 12 |
| AFIDs | AFIDs-OASIS, MNI | Rater08 | 0 | 0 | 0 |
| AFIDs | AFIDs-OASIS, MNI | Rater09 | 120 | 120 | 60 |
| AFIDs | 3T-7T | Rater01 | 72 | 72 | 72 |
| AFIDs | 3T-7T | Rater02 | 120 | 120 | 120 |
| AFIDs | 3T-7T | Rater03 | 24 | 24 | 24 |
| AFIDs | 3T-7T | Rater04 | 24 | 12 | 12 |
| AFIDs | 3T-7T | Rater05 | 12 | 36 | 48 |
| AFIDs | 3T-7T | Rater06 | 0 | 12 | 0 |
| STN | SNSX | Rater A | 120 | 120 | 60 |
| STN | SNSX | Rater B | 240 | 240 | 12 |
| STN | SNSX | Rater C | 72 | 72 | 72 |
| STN | 3T-7T | Rater A | 84 | 84 | 84 |

**Supplementary Table S2.** STN model comparison across 10 candidate regression architectures under the shared preprocessing pipeline, 5 × 4-fold participant-grouped cross-validation on the 50 training participants (100 hemisphere estimates, each averaged over the five repeats). p values are Holm-Bonferroni-corrected paired Wilcoxon signed-rank tests on participant-mean ED vs. Ridge (the selected model; n = 50).

| Model | Median ED (mm) | Mean ED ± SD (mm) | Holm p vs. Ridge |
|---|---|---|---|
| Ridge | 0.848 | 0.957 ± 0.461 | — |
| BayesianRidge | 0.899 | 0.954 ± 0.458 | 0.515 |
| PLS | 0.902 | 1.002 ± 0.483 | 0.113 |
| RandomForest | 0.908 | 1.029 ± 0.505 | 0.087 |
| SVR | 0.922 | 0.995 ± 0.477 | 0.007 |
| ElasticNet | 0.926 | 0.980 ± 0.485 | 0.087 |
| Lasso | 0.965 | 1.007 ± 0.495 | 0.018 |
| XGBoost | 0.994 | 1.055 ± 0.498 | 0.003 |
| KNN | 1.020 | 1.095 ± 0.511 | <0.001 |
| MLP | 1.952 | 2.207 ± 0.954 | <0.001 |

**Supplementary Table S3.** cZI model comparison across 10 candidate regression architectures (32-fold Leave-One-Group-Out Cross-Validation; 32 participants, 64 hemisphere estimates), using the same preprocessing, model roster, and statistical methodology as Supplementary Table S2 for the primary target. P-values are Holm-Bonferroni-corrected pairwise Wilcoxon signed-rank tests on hemisphere estimates vs. ElasticNet (the selected model, lowest median ED).

| Model | Median ED (mm) | Mean ED ± SD (mm) | Holm p vs. ElasticNet |
|---|---|---|---|
| ElasticNet | 0.808 | 0.871 ± 0.400 | — |
| SVR | 0.814 | 0.927 ± 0.395 | 0.108 |
| Lasso | 0.825 | 0.881 ± 0.398 | 1.000 |
| Ridge | 0.854 | 0.873 ± 0.398 | 1.000 |
| BayesianRidge | 0.856 | 0.876 ± 0.392 | 1.000 |
| RandomForest | 0.895 | 0.980 ± 0.444 | 0.136 |
| XGBoost | 0.907 | 0.988 ± 0.475 | 0.154 |
| PLS | 0.916 | 0.927 ± 0.415 | 0.690 |
| KNN | 0.944 | 1.001 ± 0.460 | 0.027 |
| MLP | 2.362 | 3.495 ± 3.478 | <0.001 |

**Supplementary Table S4.** fct model comparison across 10 candidate regression architectures (32-fold Leave-One-Group-Out Cross-Validation; 32 participants, 64 hemisphere estimates), using the same preprocessing, model roster, and statistical methodology as Supplementary Table S2 for the primary target. P-values are Holm-Bonferroni-corrected pairwise Wilcoxon signed-rank tests on hemisphere estimates vs. RandomForest (the selected model, lowest median ED).

| Model | Median ED (mm) | Mean ED ± SD (mm) | Holm p vs. RandomForest |
|---|---|---|---|
| RandomForest | 0.770 | 0.887 ± 0.383 | — |
| BayesianRidge | 0.781 | 0.803 ± 0.372 | 0.095 |
| ElasticNet | 0.785 | 0.806 ± 0.368 | 0.156 |
| PLS | 0.788 | 0.875 ± 0.393 | 1.000 |
| Ridge | 0.805 | 0.798 ± 0.357 | 0.095 |
| KNN | 0.808 | 0.884 ± 0.385 | 1.000 |
| Lasso | 0.815 | 0.868 ± 0.375 | 1.000 |
| SVR | 0.830 | 0.844 ± 0.350 | 0.742 |

| | | | |
|---|---|---|---|
| XGBoost | 0.848 | 0.928 ± 0.393 | 0.124 |
| MLP | 2.134 | 2.778 ± 2.234 | <0.001 |

**Supplementary Table S5.** RN model comparison across 10 candidate regression architectures (31-fold Leave-One-Group-Out Cross-Validation; 31 participants, 62 hemisphere estimates), using the same preprocessing, model roster, and statistical methodology as Supplementary Table S2 for the primary target. P-values are Holm-Bonferroni-corrected pairwise Wilcoxon signed-rank tests on hemisphere estimates vs. Lasso (the selected model, lowest median ED).

| Model | Median ED (mm) | Mean ED ± SD (mm) | Holm p vs. Lasso |
|---|---|---|---|
| Lasso | 0.458 | 0.561 ± 0.281 | — |
| Ridge | 0.475 | 0.515 ± 0.258 | 0.002 |
| PLS | 0.481 | 0.552 ± 0.288 | 0.989 |
| BayesianRidge | 0.485 | 0.517 ± 0.268 | 0.010 |
| SVR | 0.490 | 0.516 ± 0.257 | 0.069 |
| ElasticNet | 0.491 | 0.543 ± 0.278 | 0.026 |
| XGBoost | 0.499 | 0.572 ± 0.275 | 0.625 |
| RandomForest | 0.517 | 0.567 ± 0.288 | 0.989 |
| KNN | 0.604 | 0.606 ± 0.296 | 0.267 |
| MLP | 1.609 | 1.938 ± 1.602 | <0.001 |

### Supplementary Figures

**a** 16-AFID protocol subset, real MNI152NLin2009bAsym anatomy

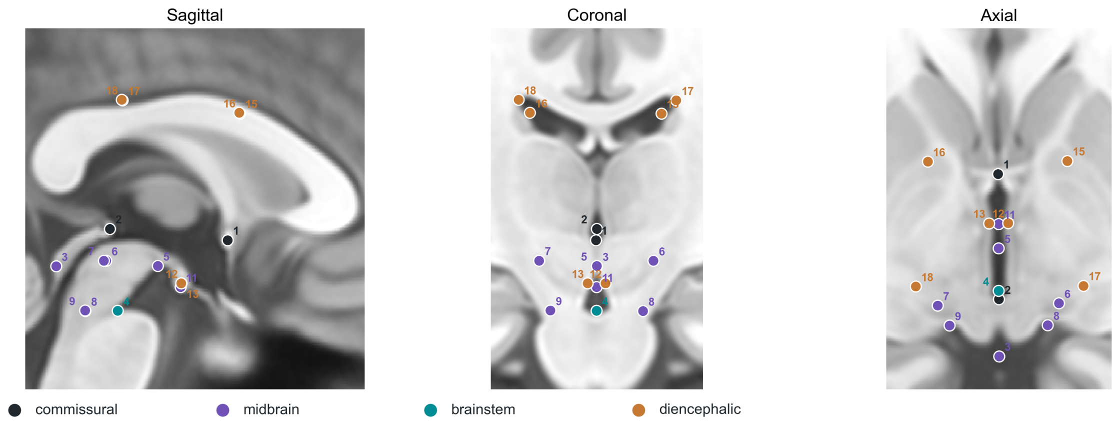


**b** Landmark index

| | | | | | |
|---|---|---|---|---|---|
| 1 | **AC** | Anterior commissure | 9 | **ILMS-L** | Left inferior lateral mesencephalic sulcus |
| 2 | **PC** | Posterior commissure | 11 | **IMS** | Intermammillary sulcus |
| 3 | **ICS** | Infracollicular sulcus | 12 | **MB-R** | Right mammillary body |
| 4 | **PMJ** | Pontomesencephalic junction | 13 | **MB-L** | Left mammillary body |
| 5 | **SIPF** | Superior interpeduncular fossa | 15 | **LVAC-R** | Right lateral ventricle at AC |
| 6 | **SLMS-R** | Right superior lateral mesencephalic sulcus | 16 | **LVAC-L** | Left lateral ventricle at AC |
| 7 | **SLMS-L** | Left superior lateral mesencephalic sulcus | 17 | **LVPC-R** | Right lateral ventricle at PC |
| 8 | **ILMS-R** | Right inferior lateral mesencephalic sulcus | 18 | **LVPC-L** | Left lateral ventricle at PC |

Positions: AFIDs ground-truth annotation on the MNI152NLin2009bAsym template (Lau et al., 2019).

**Supplementary Figure S1.** The 16-AFID protocol subset used in this study. (a) Sagittal, coronal and axial views of the MNI152NLin2009bAsym template with the 16 retained AFIDs, numbered by their AFIDs protocol index and colored by anatomical group (commissural, midbrain, brainstem, diencephalic). Points not on a displayed plane are projected onto it. (b) Protocol number, abbreviation and full name of each landmark. Positions are the AFIDs ground-truth annotation on the template (Lau et al., 2019).

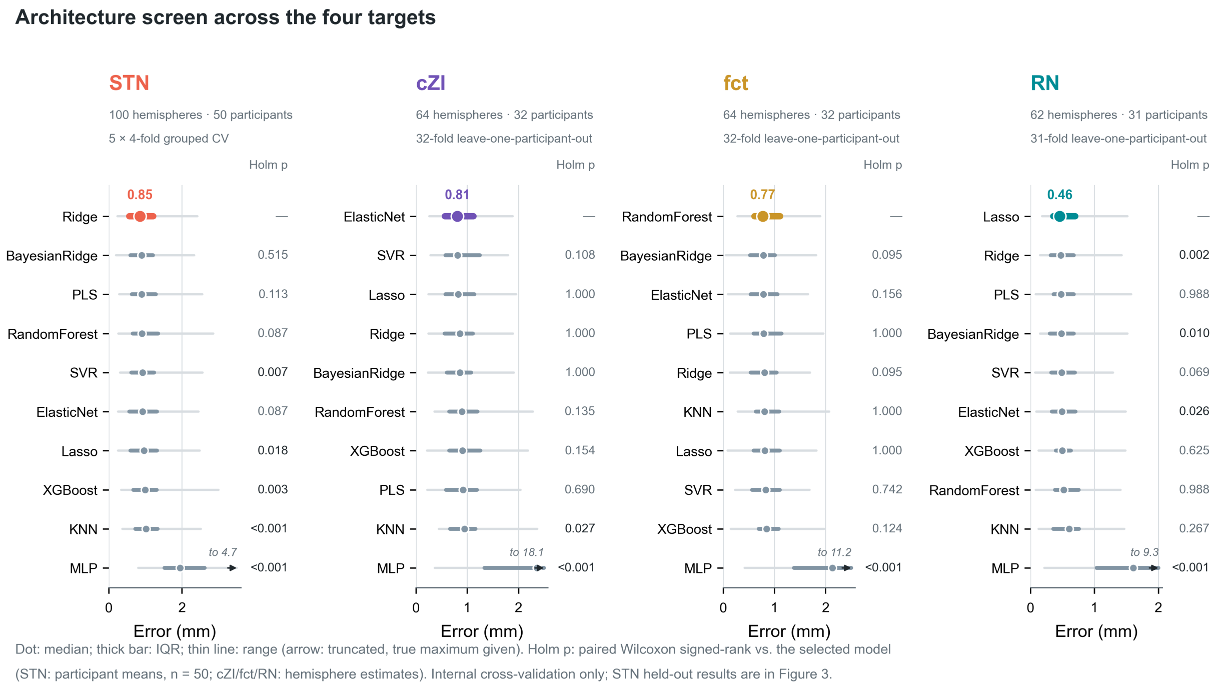


**Supplementary Figure S2.** Model comparison for all four coordinate-to-coordinate targets. For each target (subthalamic nucleus [STN], caudal zona incerta [cZI], fasciculus cerebellothalamicus [fct], and red nucleus [RN]), ten candidate regression models were screened with the same preprocessing pipeline and model roster (Supplementary Tables S2–S5). Each row shows one model's median Euclidean distance (dot), interquartile range (thick segment) and full range (thin line) over hemisphere estimates; the selected model (lowest median ED) is drawn in the target's color and its median is printed above it. Ranges extending beyond the plotted axis (MLP) are truncated with an arrow and their true maximum is given. STN: 100 hemisphere estimates from 50 participants, each averaged over 5 repeats of 4-fold participant-grouped cross-validation. cZI and fct: 64 hemisphere estimates from 32 participants (32-fold leave-one-participant-out). RN: 62 hemisphere estimates from 31 participants (31 folds). Holm p: Holm-Bonferroni-corrected paired Wilcoxon signed-rank tests against the selected model, computed on participant means for STN (n = 50) and on hemisphere estimates for cZI, fct and RN. These are internal cross-validation results; the STN held-out test is reported in main-text Figure 3.

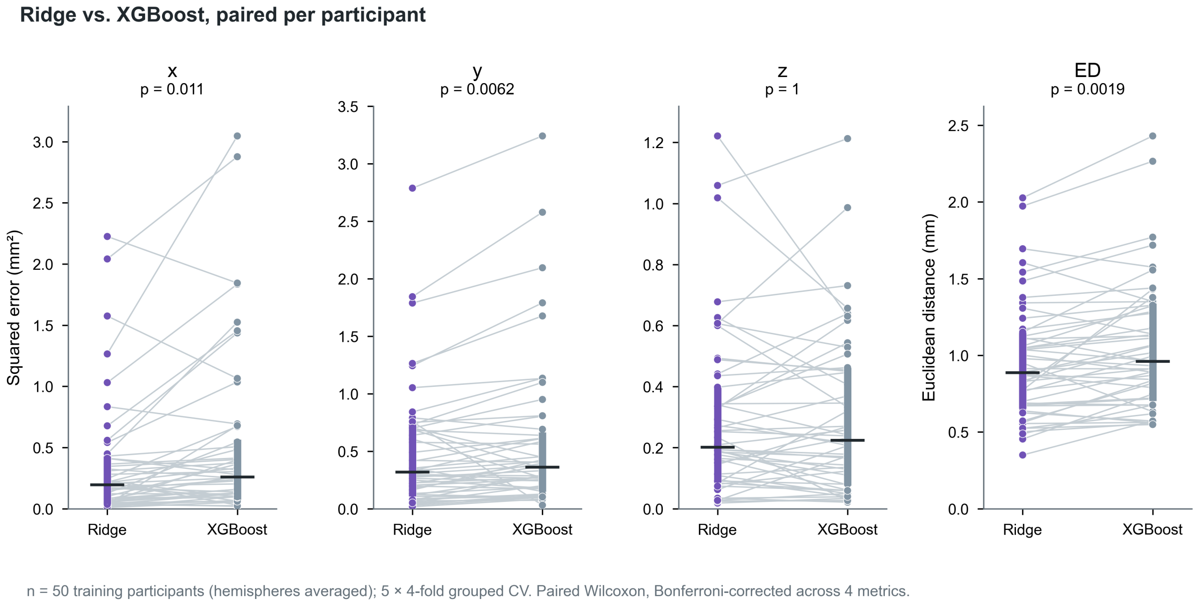


**Supplementary Figure S3.** Ridge regression versus XGBoost for the primary STN model, paired per participant. Squared error along x, y and z and Euclidean distance (ED) for each of the 50 training participants (hemispheres averaged; out-of-fold predictions, 5 × 4-fold participant-grouped cross-validation). Lines connect a participant's two models; thick bars show the IQR and ticks the median. p values are paired Wilcoxon signed-rank tests, Bonferroni corrected across the four metrics.

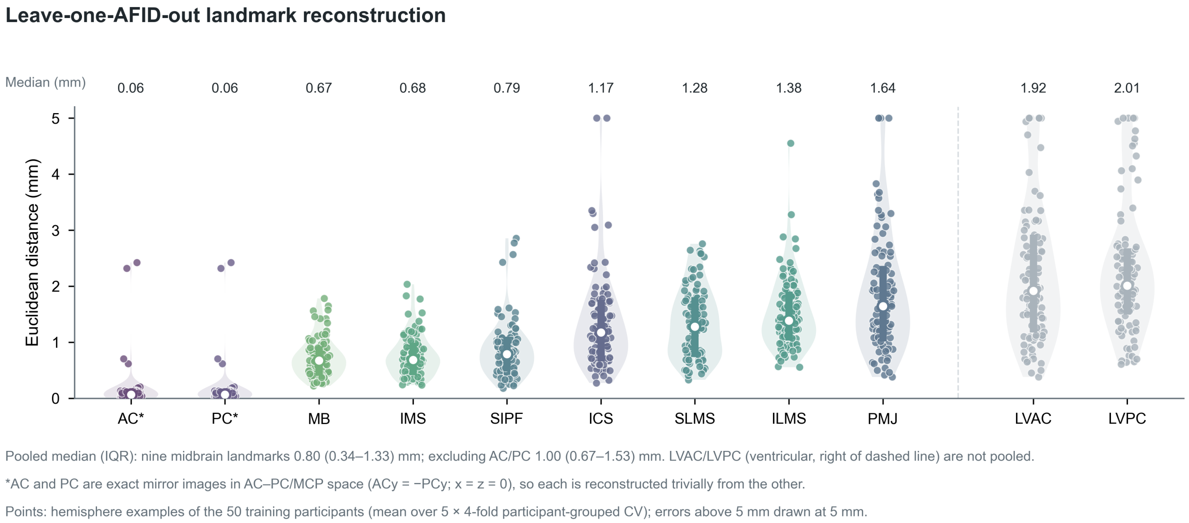


**Supplementary Figure S4.** Leave-one-AFID-out landmark reconstruction. Each of the 11 encoded landmarks was predicted (x, y, z) from the remaining ten with the STN pipeline (StandardScaler → PCA 99% → Ridge), using 5 × 4-fold cross-validation grouped by participant on the 50 training participants. Points are the 100 hemisphere examples (ED averaged over repeats); shapes are kernel density estimates, thick bars the IQR and open circles the median, which is also printed above each landmark. Landmarks are ordered by median error; LVAC and LVPC (ventricular, right of the dashed line) are excluded from the pooled midbrain summary (nine landmarks: median 0.80 mm, IQR 0.34–1.33 mm; seven landmarks excluding AC and PC: median 1.00 mm, IQR 0.67–1.53 mm). *AC and PC are exact mirror images in AC–PC/MCP space (ACy = −PCy; x = z = 0), so each is reconstructed trivially from the other. Errors above 5 mm are drawn at 5 mm.

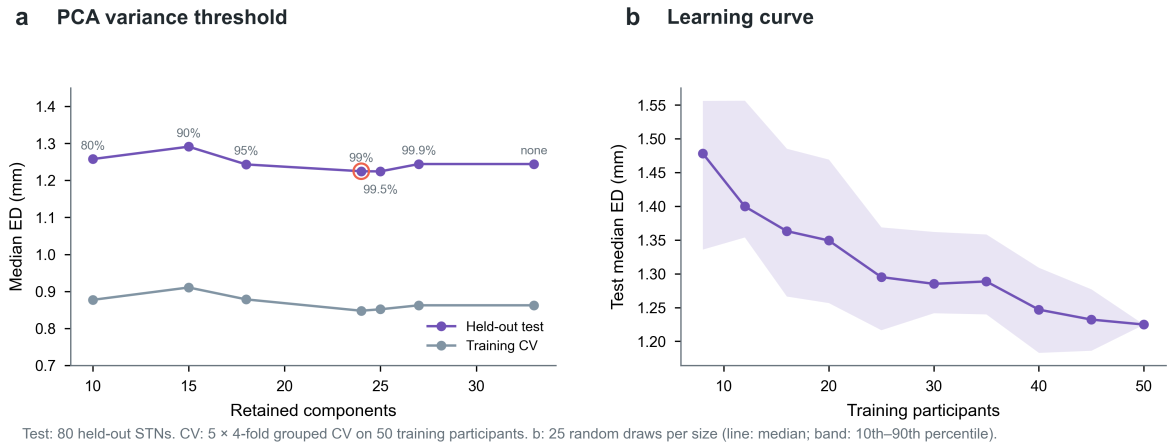


**Supplementary Figure S5.** Sensitivity of the STN pipeline. (a) Median ED against the number of retained principal components, for PCA thresholds of 80–99.9% and without PCA. The production threshold (99%) is circled. (b) Held-out median ED against the number of training participants (25 random draws per size; line, median over draws; band, 10th–90th percentile). Test values use the 80 held-out STNs.

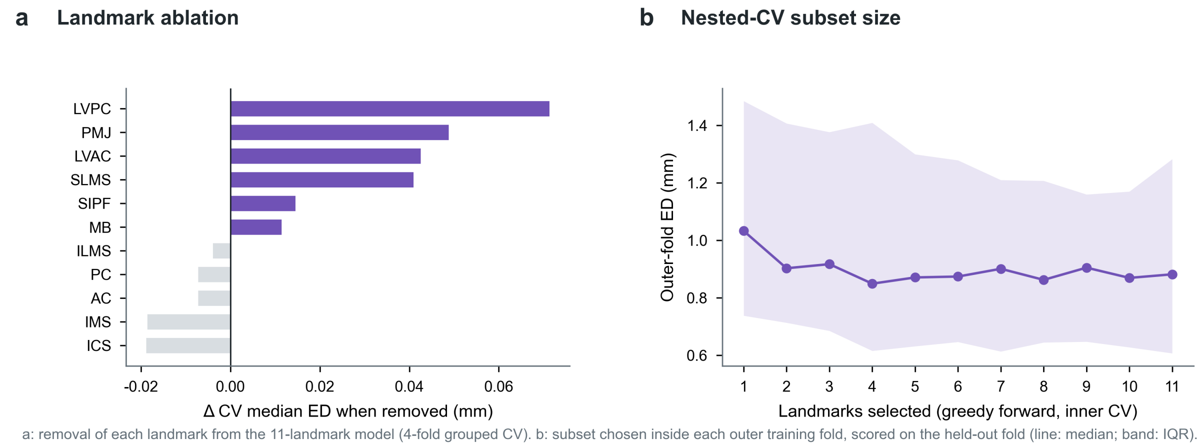


**Supplementary Figure S6.** Landmark importance for STN prediction. (a) Change in cross-validated median ED when each of the 11 encoded landmarks is removed and the pipeline refit (positive values indicate the landmark helps). (b) Outer-fold ED against subset size when greedy forward selection is nested inside cross-validation (line, median; band, IQR). Selection uses only the outer training fold, so the curve is not biased by selection.

### Supplementary Attachments

**Attachment A.** Interactive HTML quality-control reports for AFID and STN annotations, with accompanying ground-truth coordinate files (.fcsv); available at https://github.com/afids/afids-pred (data/QC).

**Attachment B.** Interactive three-dimensional visualization of all AFID and target coordinates across datasets (HTML); available at https://github.com/afids/afids-pred (figures/all_coordinates_interactive.html).

**Attachment C.** Placement protocol for the 16 AFIDs used by AFIDs-Pred, including the input file format accepted by the web tool (DOCX/PDF).
