## Supplementary Attachment C for "A coordinate-to-coordinate machine learning framework for contrast-agnostic brain target localization"

**16-AFID placement protocol for AFIDs-Pred**

### C1. Purpose and scope

This attachment specifies the subset of 16 anatomical fiducials (AFIDs) required as input to the AFIDs-Pred target-prediction models (STN, cZI, fct, RN) and the file format accepted by the deployed tool (stereotaxy.afids.io). It is a condensed, task-specific version of the full 32-point AFIDs human protocol (Lau et al., 2019; https://afids.github.io/afids-protocol), which remains the authoritative reference for landmark definitions, rater training, and worked examples. The 16 AFIDs circumscribe the midbrain and diencephalon and were selected because they surround the targets and are reliably localized (Abbass et al., 2022; Lau et al., 2019).

### C2. Before placement

- Software: 3D Slicer (v4.10 or later). The protocol was validated in v4.10.
- Images: any T1-weighted (with or without gadolinium) or T2-weighted 3D acquisition on which the midbrain sulci and the commissures are visible; the method itself is contrast-agnostic.
- Orientation: apply a rigid (linear) transform so that the volume is approximately AC–PC aligned before placing points, as described in the protocol's Linear Transforms page. This makes the midline and the AC/PC coronal planes straightforward to navigate.
- Use the full 32-point AFIDs template markups file so that each point keeps its protocol number and label.

### C3. Landmarks

Place the landmarks in the order below. AC and PC are placed first because they define the coronal planes used for LVAC/LVPC. Midline landmarks should lie on the mid-sagittal plane. Supplementary Figure S1 shows each point on the MNI152NLin2009bAsym template.

| # | Abbrev. | Landmark | Group | Placement guidance |
|---|---|---|---|---|
| **1** | AC | Anterior commissure | Commissural | Midline. Center of the anterior commissure on the mid-sagittal slice; confirm in axial and coronal views. |
| **2** | PC | Posterior commissure | Commissural | Midline. Center of the posterior commissure on the mid-sagittal slice; confirm in axial and coronal views. |
| **3** | ICS | Infracollicular sulcus | Midbrain | Midline. Inferior end of the intercollicular sulcus, where the two inferior colliculi meet at the midline. |
| **4** | PMJ | Pontomesencephalic junction | Brainstem | Midline. Ventral junction of midbrain and pons. The junction tapers gradually; use sagittal and coronal views to place the point on the ventral (pontine) side of the transition. |
| **5** | SIPF | Superior interpeduncular fossa | Midbrain | Midline. Most superior axial slice on which the interpeduncular fossa is visible; refine in the coronal view at the CSF–brain boundary below the third ventricle. |
| **6** | RSLMS | Right superior lateral mesencephalic sulcus | Midbrain | Locate on axial slices at the CSF–brain boundary; refine in coronal and sagittal views so the point sits in the sulcal angle in all three planes. |
| **7** | LSLMS | Left superior lateral mesencephalic sulcus | Midbrain | As for RSLMS, left side. |
| **8** | RILMS | Right inferior lateral mesencephalic sulcus | Midbrain | Locate on axial slices at the midbrain–pons junction; refine in the sagittal view at the change in brainstem angle at the level of the PMJ. |
| **9** | LILMS | Left inferior lateral mesencephalic sulcus | Midbrain | As for RILMS, left side. |

| | | | | |
|---|---|---|---|---|
| **11** | IMS | Intermammillary sulcus | Midbrain | Midline. Midpoint between the mammillary bodies at the gray-matter border; place in the sagittal view and refine in axial and coronal views. |
| **12** | RMB | Right mammillary body | Diencephalic | Center of the right mammillary body. |
| **13** | LMB | Left mammillary body | Diencephalic | Center of the left mammillary body. |
| **15** | RLVAC | Right lateral ventricle at AC | Diencephalic | Lateral-most aspect of the right frontal horn on the coronal slice through the AC. |
| **16** | LLVAC | Left lateral ventricle at AC | Diencephalic | As for RLVAC, left side. |
| **17** | RLVPC | Right lateral ventricle at PC | Diencephalic | Lateral-most aspect of the right lateral ventricle on the coronal slice through the PC. |
| **18** | LLVPC | Left lateral ventricle at PC | Diencephalic | As for RLVPC, left side. |

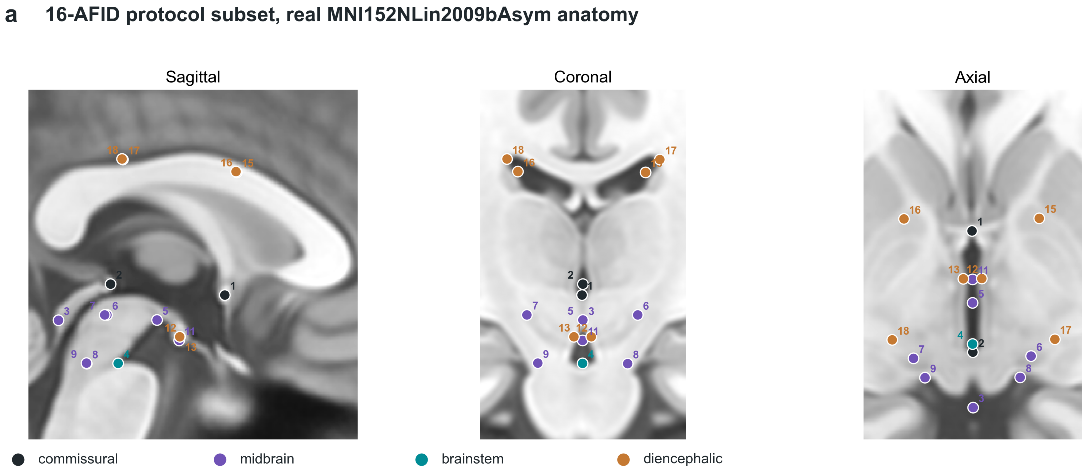


**b Landmark index**

| | | | | | |
|---|---|---|---|---|---|
| 1 | **AC** | Anterior commissure | 9 | **ILMS-L** | Left inferior lateral mesencephalic sulcus |
| 2 | **PC** | Posterior commissure | 11 | **IMS** | Intermammillary sulcus |
| 3 | **ICS** | Infracollicular sulcus | 12 | **MB-R** | Right mammillary body |
| 4 | **PMJ** | Pontomesencephalic junction | 13 | **MB-L** | Left mammillary body |
| 5 | **SIPF** | Superior interpeduncular fossa | 15 | **LVAC-R** | Right lateral ventricle at AC |
| 6 | **SLMS-R** | Right superior lateral mesencephalic sulcus | 16 | **LVAC-L** | Left lateral ventricle at AC |
| 7 | **SLMS-L** | Left superior lateral mesencephalic sulcus | 17 | **LVPC-R** | Right lateral ventricle at PC |
| 8 | **ILMS-R** | Right inferior lateral mesencephalic sulcus | 18 | **LVPC-L** | Left lateral ventricle at PC |

Positions: AFIDs ground-truth annotation on the MNI152NLin2009bAsym template (Lau et al., 2019).

**Figure C1.** The 16 AFIDs used by AFIDs-Pred on the MNI152NLin2009bAsym template (reproduced from Supplementary Figure S1).

### C4. Export and file format

- Export the markups as a Slicer fiducial file (.fcsv) with the standard three header lines.
- The file must contain all 32 rows of the AFIDs template in protocol order (1–32). Prediction reads points by position, not by label. The 16 points that are not used (protocol #10, 14 and 19–32) may be left at any coordinate but must not be deleted. Deleting a row shifts every subsequent point and silently corrupts the input.

- Coordinates are RAS millimeters in native image space. The web tool derives the AC–PC transformation from the uploaded AFIDs and returns the predicted STN coordinates together with that transformation matrix. The prediction code in the afids-pred repository operates on AC–PC-aligned, MCP-centered AFID coordinates.
- The web tool currently returns STN predictions only. The cZI, fct and RN models are distributed as model files in the afids-pred repository.

### C5. Quality control

- Verify that every midline point (AC, PC, ICS, PMJ, SIPF, IMS) lies on the mid-sagittal plane after AC–PC alignment. A lateral offset of more than about 1 mm usually indicates a misplaced point or an unhardened transform.
- Check that each bilateral pair (SLMS, ILMS, MB, LVAC, LVPC) is roughly symmetric about the midline.
- Check the AC–PC distance. Across the 202 scans used in this study it had a median of 26.3 mm (range 22.5–31.8 mm); values far outside this range suggest an AC or PC error. Both commissures are inputs to the model and also define the coordinate frame.
- Prediction error grows with AFID placement error: approximately 0.46 mm of additional target error per 1 mm of mean AFID error (main text, section 4.4 and Figure 4b). Re-check any landmark that is not clearly visible.
- Example QC reports for the datasets used in this study are provided in Supplementary Attachment A (https://github.com/afids/afids-pred, data/QC).

### C6. Intended use

AFIDs-Pred is a research tool. Its output is a predicted coordinate for the centroid of the target structure and is not a substitute for direct visualization, microelectrode recording, or clinical judgement. Predictions for cZI, fct and RN were validated only by cross-validation in healthy controls (Supplementary Methods S1).
